# NMA: Network meta-analysis based on multivariate meta-analysis and meta-regression models in R

**DOI:** 10.1101/2025.09.15.25335823

**Authors:** Hisashi Noma, Kazushi Maruo, Shiro Tanaka, Toshi A. Furukawa

## Abstract

Network meta-analysis has become an established methodology within systematic reviews for comparing the effectiveness of multiple treatments, and it has been now a standard approach in comparative effectiveness research. However, the underlying statistical methods are often highly technical for non-statisticians in practice, and no freely available software package has been developed that can handle a general framework based on the multivariate meta-analysis and meta-regression models. To address these issues, we developed `NMA`, a comprehensive and user-friendly R package that covers extensive analysis and graphical tools of network meta-analysis with simple commands. The `NMA` package provides generic functional tools for evidence synthesis based on the multivariate meta-analysis models, network meta-regression, assessment of heterogeneity and inconsistency, comparative effectiveness analyses, and a range of graphical tools. In addition, `NMA` includes data-handling functions that facilitate the integration of both arm-level data and summary effect measure statistics easily. In this article, we provide a gentle introduction to the `NMA` package and illustrate its application through a case study of a network meta-analysis of antihypertensive drugs.

**Highlights:** *What is already known?*

- Several freely computational packages are available for network meta-analysis, but no general frequentist tool based on the multivariate meta-analysis and meta-regression models, introduced by White et al. ^13^, has been developed.

*What is new?*

- We developed `NMA`, a comprehensive R package for network meta-analysis based on multivariate meta-analysis and meta-regression models with frequentist approach.
- The `NMA` package provides a broad range of functions for evidence synthesis, heterogeneity and inconsistency assessment, comparative effectiveness analysis, and graphical visualization.
- Key analytical tools—such as Higgins’ global inconsistency test ^12^, network meta-regression, and advanced inferential and prediction methods to address invalidity issues of the ordinary approaches ^17,21^ —are fully implemented.
- Generic data-handling tools can now integrate arm-level data with summary statistics. This provides greater flexibility in network meta-analysis, making it especially useful for studies of survival outcomes.

*Potential impact for RSM readers*

- The package facilitates the practical use of network meta-analysis for a broad range of researchers, including non-statisticians, thereby enhancing the accessibility of systematic reviews on important clinical and public health questions.
- By comprehensively covering standard analyses and graphical tools, the `NMA` package is also valuable for educational purposes, serving as a practical resource for students, researchers, and clinicians to learn the research methods through real-world case studies.

## 1. Introduction

In almost all disease areas today, multiple treatment options exist, and the question of which treatment constitutes the best option for each patient has become a central issue ^1^. At the same time, with the profound impact of population aging in developed countries, this question has also become increasingly important from the perspectives of health economics and public health policy ^2,3^. Conventional pairwise meta-analysis, which has been developed since the 1980s, has substantial limitations for addressing this purpose, and network meta-analysis has become the new standard method for comparative effectiveness research ^4,5^. Naci and O’Connor ^6^ have suggested that network meta-analysis can also be applied at the stage of regulatory approval of new drugs, in order to compare their efficacy and safety with those of existing therapies. The necessity of such approaches is expected to increase substantially in the future.

Several excellent free computational tools for network meta-analysis have already been developed in R (R Foundation for Statistical Computing, Vienna, Austria). For Bayesian approaches, which have been widely adopted since their early development, `gets` ^7^, `pcnetmeta` ^8^ and `multinma` ^9^ packages are available. For frequentist approaches, the contrast-based approach is comprehensively supported by the `netmeta` package ^10^. The `metafor` package ^11^ also provides extensive functionalities that are highly useful for conducting complex integrated analyses, including network meta-analysis. While each of these packages implements sophisticated functions reflecting the latest methodological developments, the frequentist framework founded on the multivariate meta-regression framework introduced by Higgins et al. ^12^ and White et al. ^13^ has been one of the standard methodological framework of network meta-analysis. In Stata, the `network` package ^14^ provides a general computational tool for this framework, but no such tool has been available in R. In addition, none of the current network meta-analysis software accommodates methods to overcome fundamental invalidity problems in the inferences of treatment effects, where confidence intervals obtained by standard methods can severely underestimate statistical error ^15–17^.

To address these gaps, we have developed the R package `NMA` as an extensive computational tool for practice of network meta-analysis, designed so that all functions can be easily used by non-statisticians. In addition to the functions of Stata’s `network` package, the `NMA` package implements various methods based on multivariate meta-regression models, such as Jackson’s random inconsistency model ^18,19^ and the multivariate *I*² statistic ^20^. Furthermore, it incorporates new inference and prediction methods for treatment effects based on higher-order asymptotic theory. ^17,21^ A wide range of graphical tools have also been implemented, including those for assessing transitivity ^22^. The package provides enhanced data-handling functionality, with tools that allow integration of both arm-level data and summary-level data. This provides greater flexibility in network meta-analysis, making it especially useful for studies of survival outcomes.

In this paper, we provide an overview of the functions implemented in the `NMA` package and illustrate its use with example programs and case analyses. In Section 2, we briefly describe the example data used in this article. Section 3 presents a summary of the functions implemented in the `NMA` package, and Sections 4 through 8 demonstrate various applications of the package using the network meta-analysis of antihypertensive drugs reported by Sciarretta et al. ^23^ as a case study with binary outcome. Additional analytical tools that cannot be fully described in the main text are detailed in the Supplementary Materials.

Due to space limitations, the R output has been selectively presented. Complete R programs reproducing all case analyses are available on the authors’ GitHub page (https://github.com/nomahi/NMA). In addition, further examples with continuous and survival outcomes are given in e-Appendices A and B, respectively.

## 2. Illustrative example: Network meta-analysis of antihypertensive drugs

In this article, we present a case analysis of `NMA` using the network meta-analysis of antihypertensive drugs reported by Sciarretta et al. ^23^ as an example of a binary outcome. Sciarretta et al. ^23^ conducted a network meta-analysis comparing seven classes of antihypertensive drugs—α-blockers (AB), angiotensin-converting enzyme inhibitors (ACEIs), angiotensin II receptor blockers (ARBs), β-blockers (BBs), calcium channel blockers (CCBs), conventional treatment (CT), and diuretics (DDs)—with placebo. The outcome of interest was the occurrence of heart failure, treated as a binary variable. The dataset included 26 randomized controlled trials conducted between 1997 and 2009, comprising a total of 223,313 participants.

Within the `NMA` package, this is provided as a dataset `heatfailure` and contains arm-level data, including sample sizes (`n`) and event counts (`d`) for each group. In addition, arm-level covariate data are available, including mean baseline systolic and diastolic blood pressure (`SBP, DBP`), as well as year of publication (`pubyear`).

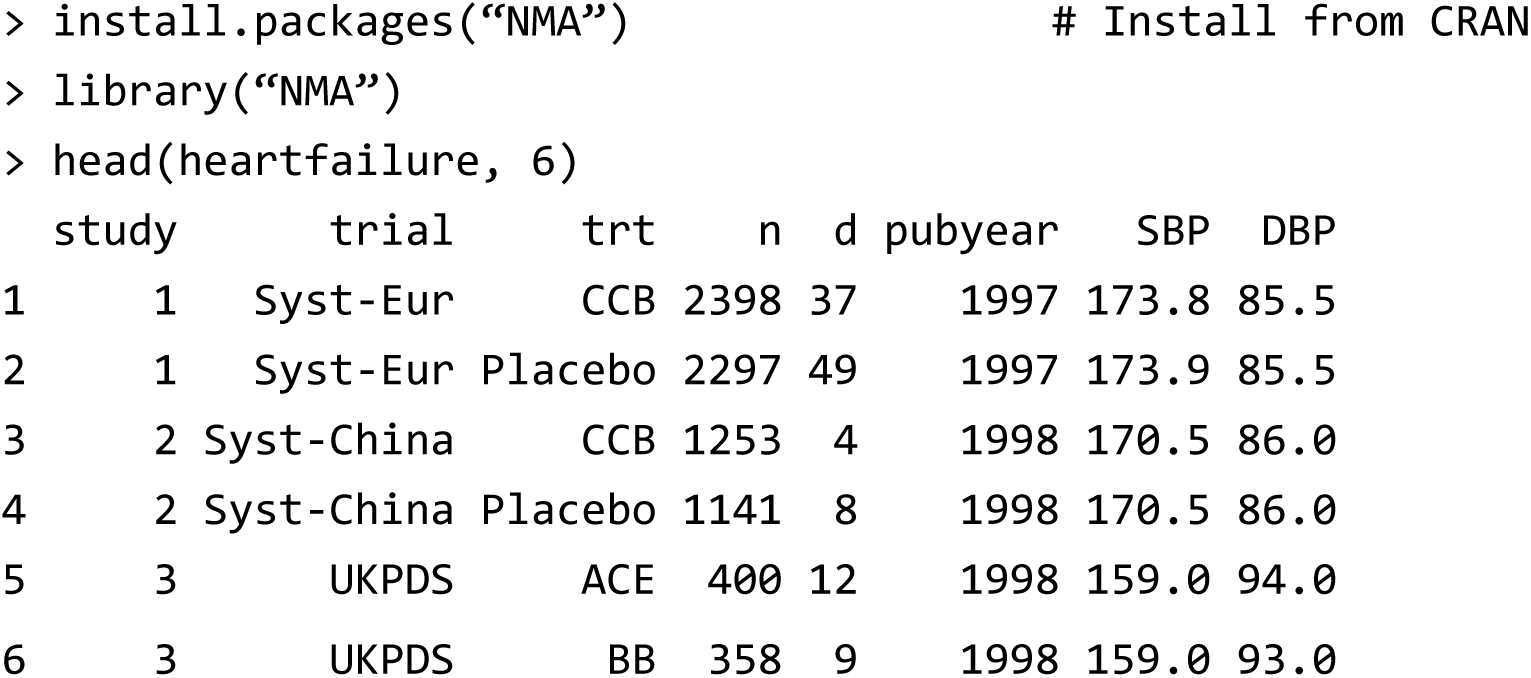

## 3. General overview of the NMA package

### 3.1 Functionalities and features

The R package `NMA` was developed as a collection of generic computational tools for network meta-analysis. It is based on the multivariate meta-regression framework proposed by White et al. ^13^ and is designed to apply effective statistical methods available within this model. Table 1 provides an overview of the functions implemented in the package.

**Table 1.** Main functions of the NMA Package and their descriptions.

| Function | Description | Section |
| --- | --- | --- |
| `setup` | Transforming arm-level data into contrast-based summary statistics and creating data objects for all subsequent analyses. | 3.2 |
| `trans.armdata` | Transforming reported contrast-based summary statistics into arm-level data. | 3.2 |
| `trans.armdataP` | Transforming reported incidence proportion or survival probability data into arm-level data. | 3.2 |
| `netplot` | Generating a network diagram. | 4.1 |
| `SumPMA` | Pairwise meta-analyses for all directly compared treatment pairs in the network. | 4.2 |
| `nma` | Performs a network meta-analysis using a multivariate meta-analysis model and computes heterogeneity measures. | 5.3 |
| `nmareg` | Performs a network meta-regression using a contrast-based approach. | 5.3 |
| `nmarank` | Calculates ranking statistics for the network meta-analysis. | 6.1 |
| `nmaleague` | Generates a league table. | 6.2 |
| `nmaforest` | Generates a ranked forest plot of the network meta-analysis results. | 6.3 |
| `local.ict` | Local inconsistency tests for all closed loops in the network. | 7.1 |
| `global.ict` | Higgins' global inconsistency test. | 7.2 |
| `nmaQ` | Computes the multivariate Q-statistic and its factorization. | 7.2 |
| `sidesplit` | Conducts sidesplitting to quantify direct and indirect evidence for all possible treatment pairs and to test inconsistency. | 7.3 |
| `random.icm` | Jackson's random inconsistency model. | 7.4 |
| `nmafunnel` | Generates a comparison-adjusted funnel plot. | 8.1 |
| `nmaweight` | Evaluates study weights and the contribution matrix. | 8.2 |
| `transitivity` | Assesses transitivity. | 8.3 |

The package incorporates numerous features that have not been available in existing R packages, including network meta-regression, Higgins’s global inconsistency test ^12^, Jackson’s random inconsistency model ^18^, inference and prediction methods based on higher-order asymptotics developed by Noma et al. ^17,21^, and tools for assessing transitivity. In addition, the package offers enhanced data-handling functionality, implementing editing tools that allow the seamless integration of arm-based data (e.g., means, standard deviations) with trials reporting only contrast-based effect measures (e.g., mean differences).

To ensure accessibility for non-statisticians, the package has been designed with a user-friendly interface. Once an analysis dataset is generated using the `setup` function, all analytical tools can be applied with simple commands using the generated object. In the following sections, we provide an overview of the purpose of each function, how to use them, and how to interpret their results.

### 3.2 Supported data types and setup

The `NMA` package accommodates three types of outcomes—continuous, binary, and survival—as summarized in Table 2. For each outcome type, the package enables estimation using several effect measures, including the mean difference, standardized mean difference, risk difference, risk ratio, odds ratio, hazard ratio, and survival probability difference.

**Table 2.** Outcome types supported in the NMA package and their descriptions.

| Type of outcome | Effect measure | Description |
| --- | --- | --- |
| Continuous | Mean difference | For the included trials, arm-level data on the mean, standard deviation, and sample size are required. If some trials report only summary statistics, the `trans.armdata` function can be used to back-calculate the corresponding arm-level data (see e-Appendix A). |
|  | Standardized mean difference |  |
| Binary | Risk difference | For the included trials, arm-level data on the number of events, and sample size are required. If some trials report only summary statistics, the `trans.armdata` function can be used to back-calculate the corresponding arm-level data (see Section 3.2). |
|  | Risk ratio |  |
|  | Odds ratio |  |
| Survival | Hazard ratio | For the included trials, either the estimated log-hazard ratio with its standard error or arm-level data on the number of events and sample size are required. For convenience, the `trans.armdata` function can be used to convert the estimated log-hazard ratio, under the proportional hazards assumption, into arm-level data on the number of events and sample size that carry equivalent statistical information. Also, the arm-level data can be estimated from survival probability estimates using the `trans.armdataP` function. When the analysis is performed using the `setup` function, the original hazard ratio estimates are restored, enabling all subsequent analyses to be conducted (see e-Appendix B). |
|  | Survival probability difference |  |

All analyses require that data be prepared in *arm-level* format. These arm-level datasets are edited by the `setup` function, which edits and creates contrast-based statistics suitable for the analyses using multivariate meta-analysis models. As in conventional meta-analysis, however, mixed data of arm-level and summary-level statistics are often created in practice ^24^. The `NMA` package therefore provides dedicated functions for handling such cases. For example, in continuous outcomes, some trials may report mean, standard deviation, and sample size for each arm, whereas others provide only contrast-based effect measures such as the mean difference with a standard error (which can be reconstructed from confidence intervals or *p*-values). In the latter case, the function `trans.armdata` can be used to back-calculate (pseudo) arm-level data. Note that the reconstructed values will not necessarily reproduce the original arm-level data exactly. However, in practice, the `setup` function transforms all the (pseudo) arm-level data into contrast-based effect measures for analysis, and the resulting summary measures are designed to coincide closely with the originally reported effect measures. Thus, the arm-level data generated by `trans.armdata` should be viewed as “working” pseudo-data, serving as a bridge to construct contrast-based summaries.

For survival outcomes, hazard ratios are frequently reported ^25^. These summary data can also be passed to `trans.armdata`. Under the proportional hazards assumption, the function applies the complementary log–log transformation to invert the hazard ratio formula ^26^ ^27^, thereby generating pseudo-data in the form of reconstructed event counts. Also, arm-specific survival probability data can sometimes be obtained from a Kaplan–Meier plot ^25^. These survival probabilities can be converted into (pseudo) event counts and sample sizes using the `trans.armdataP` function. These pseudo-data can then be combined with trials reporting only event counts and sample sizes, producing a dichotomized arm-level dataset across all studies. Passing this dataset to the `setup` function yields contrast-based effect measures expressed as hazard ratios, which reproduce the original estimates.

Once arm-level data from different-type data are merged into a single dataset, the `setup` function can produce contrast-based effect measures. All subsequent analyses can then be performed using the resulting object with simple commands. If arm-level covariates are available—for example, summary baseline characteristics that can be used in meta-regression or for evaluating transitivity—these variables can be added directly to the dataset in advance. Such data editing may conveniently be carried out in spreadsheet software such as Microsoft Excel (Microsoft, Redmond, WA.).

As an illustrative example for the binary outcome case, the required variables include study identifier (`study`), treatment (`trt`), number of events (`d`), sample size (`n`), and covariates (`z`; optional, multiple allowed). Users must also specify the effect measure of interest (risk difference, risk ratio, or odds ratio) and designate a reference treatment (`ref`).

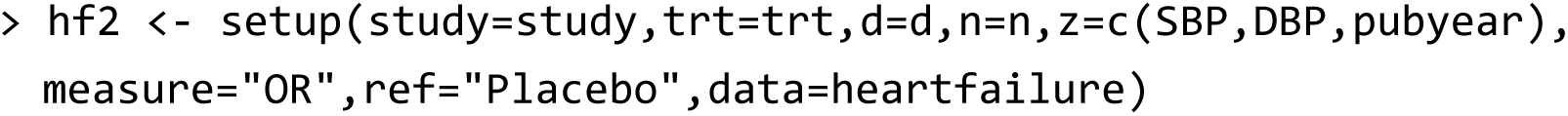

By using the output object generated by the setup function as an argument, all subsequent analyses can be executed with simple commands.

## 4. Summarizing basic information on the network of evidence

### 4.1 Network diagram

In network meta-analysis, network diagrams ^28^ are presented in most studies to visualize both the amount and structure of the evidence being synthesized. The `NMA` package facilitates this process through the `netplot` function, which enables users to generate such diagrams in a straightforward manner. The function can be executed using the syntax illustrated below, and its specifications can be flexibly modified according to user preferences. The size of the nodes and the thickness of the edges in the `netplot` function are proportional to the total sample size for each treatment and each direct comparison, respectively.

> netplot(hf2)

*(The figure created is shown in Figure 1.)*

**Figure 1.**
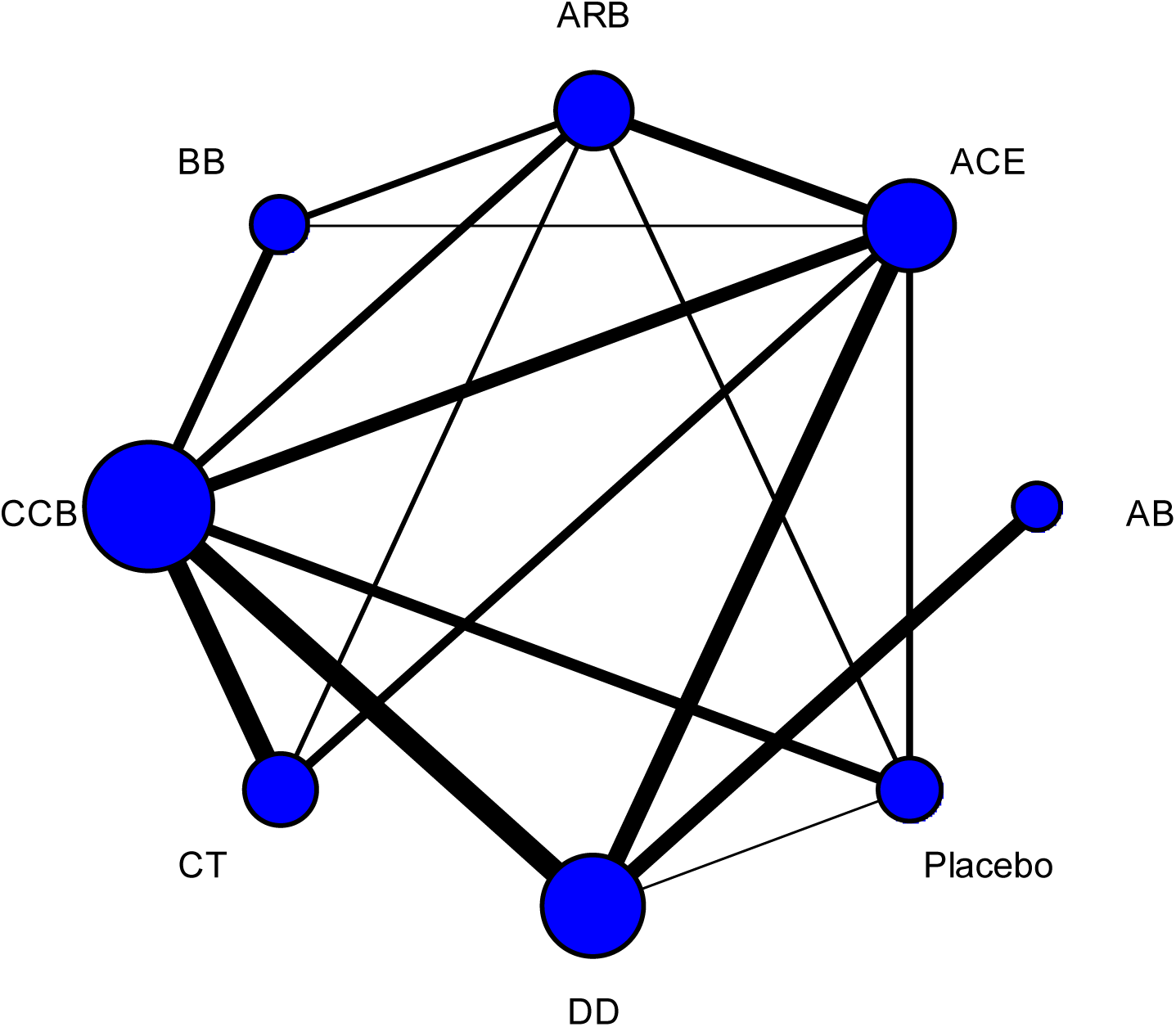
Network diagram for the antihypertensive drug study created by the `netplot` function.

### 4.2 Pairwise meta-analysis for all treatment pairs with direct comparisons

As part of the basic information, the results of pairwise meta-analyses for all direct comparisons in the network can be summarized using the `SumPMA` function. This output includes the results of meta-analyses based on the DerSimonian–Laird-type random-effects model ^29^, along with estimates of *τ*², *I*², and *H*² statistics, and the Egger test ^30^. By default, the restricted maximum likelihood (REML) method is adopted.

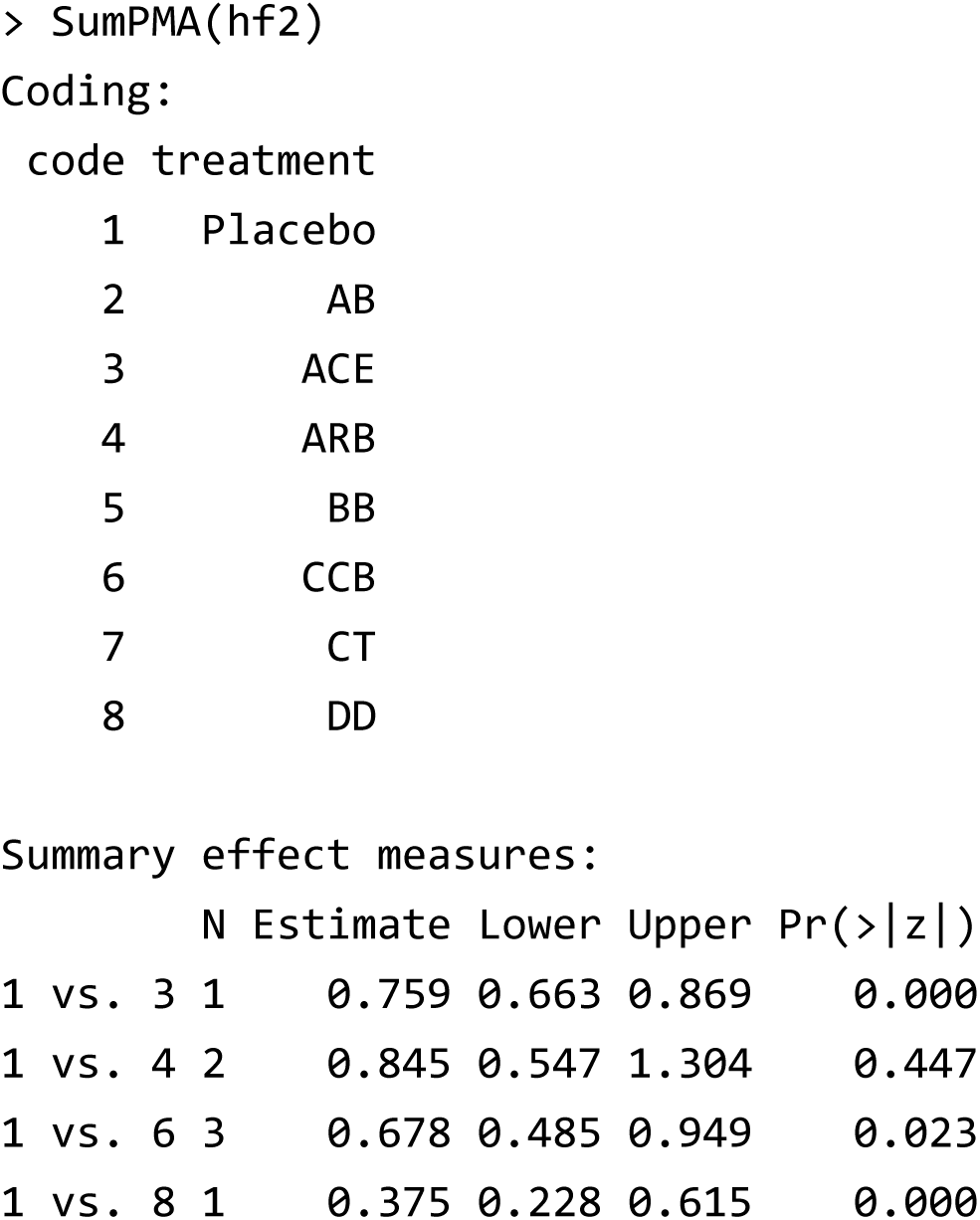

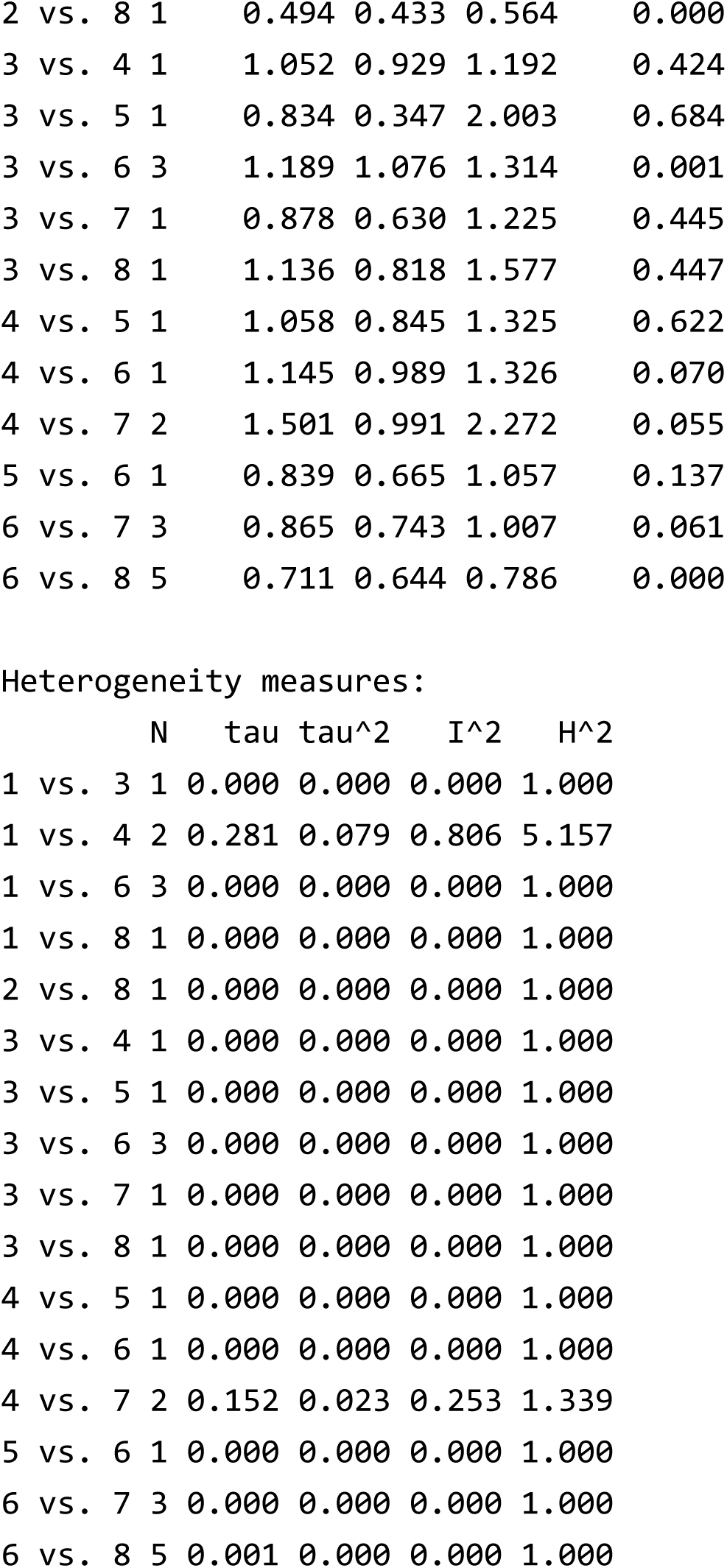

## 5. Network meta-analysis using multivariate meta-analysis models

### 5.1 Multivariate meta-analysis models and synthesis methods

The contrast-based approach is one of the principal methods used for evidence synthesis in network meta-analysis ^22,31^. In this framework, effect measures such as odds ratios, hazard ratios, or mean differences are modeled as outcome measures. This approach can be interpreted as a generalization of the conventional DerSimonian–Laird method for pairwise meta-analysis ^29^, whereby study-specific outcomes are treated as multivariate outcomes and modeled within a multivariate meta-analysis framework ^32^.

White et al. ^13^ demonstrated that such multivariate meta-regression models allow for the implementation of contrast-based network meta-analysis, and further showed that this framework provides a general means to evaluate inconsistency through the design-by-treatment interaction model ^12^. In this section, we outline the underlying statistical model and corresponding analytical procedures.

Specifically, we consider the synthesis of *N* trials comparing *p* + 1 treatments. Let *Y_ij_* denote the estimator of the treatment effect of the *j*th treatment relative to a designated reference treatment (e.g., placebo) in the *i*th trial (*i* = 1, 2, …, *N*; *j* = 1,2, …, *p*). Commonly used effect measures include the mean difference, standardized mean difference, risk difference, risk ratio, odds ratio, and hazard ratio, with ratio measures typically modeled on the logarithmic scale. For the contrast-based network meta-regression, we adopt a multivariate meta-analysis model for the outcome variable ***Y****_i_* = (*Y_i_*_1_, *Y_i_*_2_,…, *Y_ip_*)*^T^*,

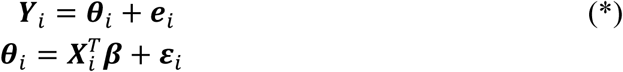

where ***θ*** = (*θ_i_*_1_, *θ_i_*_2_,…, *θ_ip_*)*^T^* and ***β*** = (***β****^T^*,…, ***β****^T^*)*^T^*. The regression function model 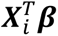 involves a design matrix,

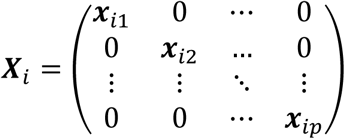

where ***x****_ij_* is a *q_j_* × 1 covaraiate vector for *Y_ij_* (*q_j_* is the number of the covariates) and ***β****_j_* is its *q_j_* × 1 regression coefficient vector. Also, ***e****_i_* and ***ε****_i_* are independent random variation terms within and across studies (*p* × 1 random vectors), assumed to be distributed ***e****_i_*∼MVN(**0**, ***S****_i_*) and ***ε****_i_*∼MVN(**0**, ***Σ***). ***S****_i_* (a *p* × *p* matrix) is the within-study covariance matrix,

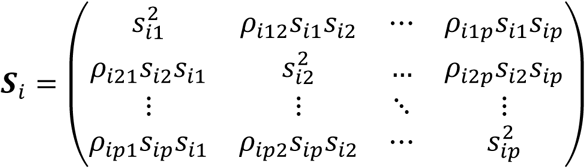

which is usually assumed to be known and fixed to its valid estimate ^33^. In addition, ***Σ*** is the between-studies variance‒covariance matrix:

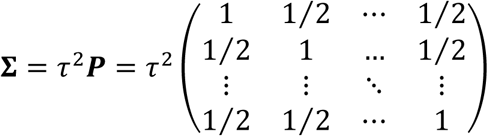

for *τ*^2^ > 0. Note that the correlation structure of ***Σ*** can be assumed to be unstructured, however, there are rarely enough studies to allow identification of all variance–covariance parameters. Consequently, most network meta-analyses adopt the equal-variance assumption for the *p* components of ***ε****_i_*. Under this assumption, and by virtue of the consistency property, all pairwise correlation coefficients are set to 0.50 ^34^. In the `NMA` package, we likewise adopt the equal-variance assumption as the standard specification of the model.

For trials that do not include a reference treatment, the data augmentation strategy proposed by White et al. ^13^ can be applied; in this approach, a quasi-negligible dataset is added to the reference arm (e.g., 0.001 events for 0.01 patients in the case of a binary outcome). In addition, because most individual clinical trials typically involve only two to four arms, we formally employ subvectors or submatrices in the likelihood function ^13^.

Standard network meta-analysis without covariates and under the consistency assumption can be expressed as a special case of the model with ***x****_ij_* = (**1**), a vector of ones. Meta-regression analyses incorporating covariates can then be performed within the same framework. In terms of the classification by Donegan et al. ^35^, the multivariate meta-regression model based on the framework proposed by White et al. ^13^ corresponds to the common interaction assumption, where the treatment-by-covariate interaction is assumed to be the same across all comparisons. It should be noted that the regression function is modeled separately for each outcome. Furthermore, when *τ*^2^ = 0, the model reduces to a fixed-effect model. In the `NMA` package, standard network meta-analyses can be conducted using the `nma` function, while meta-regression can be performed with the `nmareg` function.

Model parameters can be estimated using the standard REML method. Under a random-effects model, as in pairwise meta-analyses, the confidence intervals for ***β*** may be inaccurate when the number of studies *N* is small ^16,17,36^. The Cochrane Handbook ^24^ recommends the use of the HKSJ method ^15,37,38^ for pairwise meta-analyses. For network meta-analysis, Noma et al. ^17^ also proposed a higher-order asymptotic correction, which has been adopted as the default method in the `NMA` package.

### 5.2 Assessment of heterogeneity

The issue of between-trial heterogeneity in treatment effects is also of central importance in network meta-analysis. The conventional measures used in pairwise meta-analysis, such as Cochrane’s *Q*-test ^39^ and Higgins’ *I*^2^ statistic ^40^, have been extended to the multivariate meta-analysis framework by Jackson et al. ^20^, and can be directly computed from model (*). The parameter *τ* can be interpreted as the heterogeneity standard deviation. In addition, prediction intervals ^41^, which represent the expected range of treatment effects in future populations while accounting for between-study heterogeneity, have become widely used in recent meta-analyses. Although prediction intervals have commonly been calculated using approximations based on the *t*-distribution, concerns about their accuracy have been raised ^36,42^. To address this issue, Noma et al. ^21^ proposed several improved methods. The `NMA` package adopts, as the default method, an approach based on higher-order asymptotic approximations developed by Noma et al. ^21^, and these results are reported collectively in the output of the `nma` function.

### 5.3 Computational tools for the primary synthesis methods

The synthesis analysis based on the consistency assumption can be performed by the `nma` function, which adopts the higher-order asymptotic correction methods proposed by Noma et al. ^17,21^ as default (named as “Noma-Hamura” methods). The output also provides various heterogeneity statistics, including *τ*, *τ*^2^, the *H*^2^ statistic, the *I*^2^ statistic, the *Q*-test, and the prediction interval. By setting `eform` `=` `TRUE`, the estimated effect measures are exponentiated, thereby allowing direct interpretation of effect sizes such as odds ratios, risk ratios, and hazard ratios, which are synthesized on the logarithmic scale under normality assumptions.

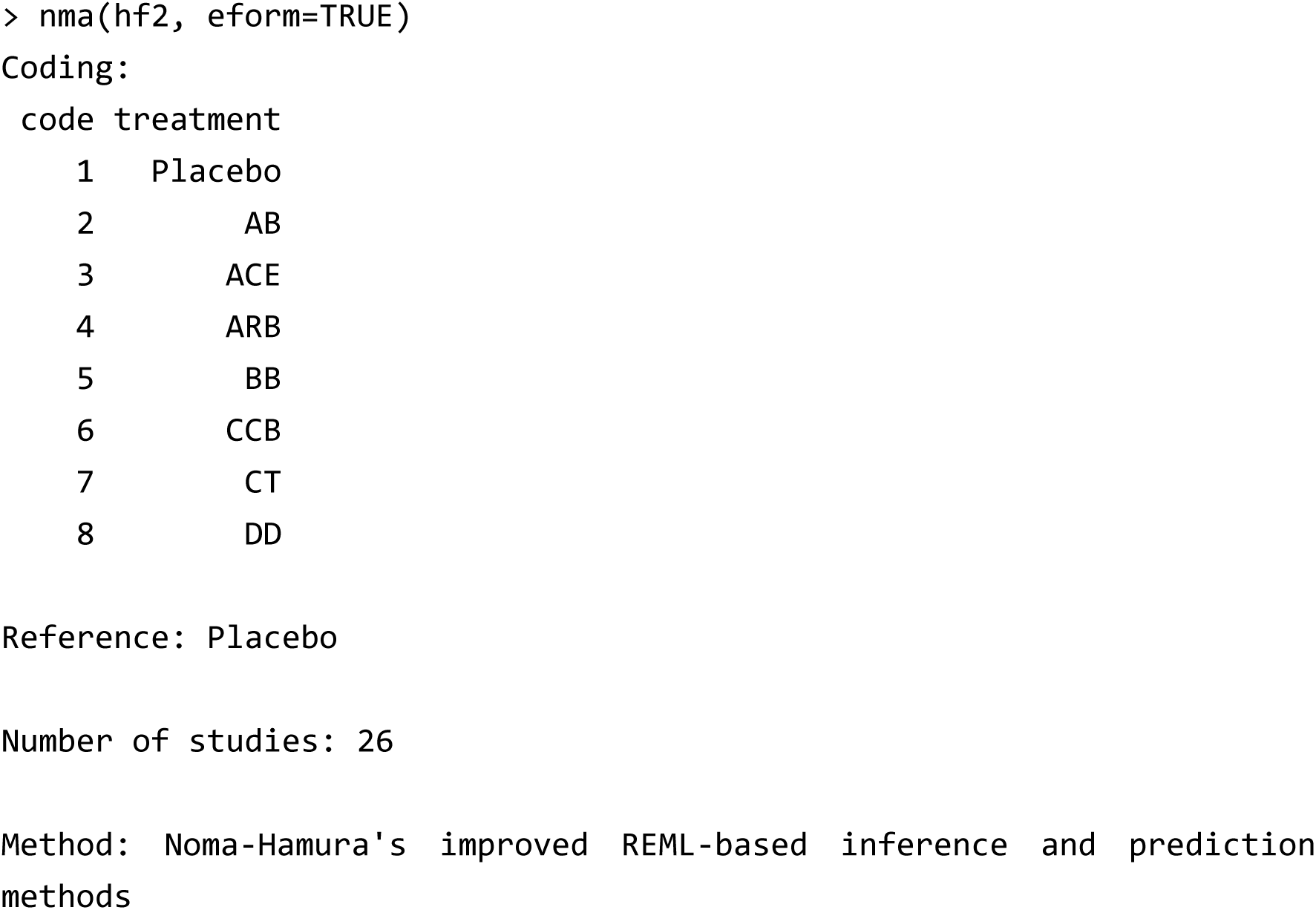

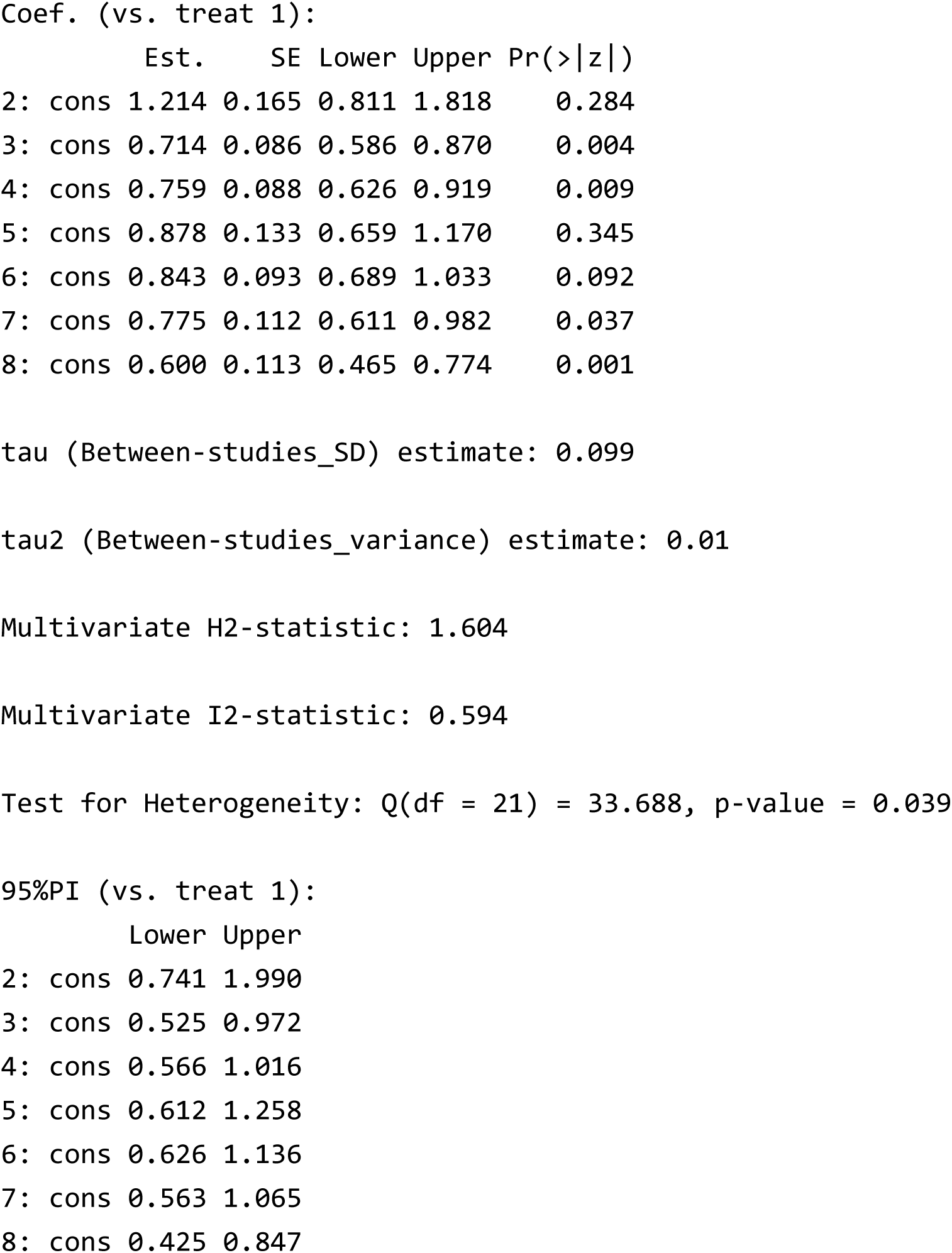

Similarly, network meta-regression can be conducted using the `nmareg` function. The example below illustrates an analysis of effect modification for the odds ratio of ACE versus placebo with systolic blood pressure (SBP) included as a covariate. It should be noted that in multivariate meta-regression, a different regression function is specified for each component of the outcome vectors ***Y***_1_,…, ***Y****_N_*. The variable to be evaluated as an effect modifier is specified via the argument `z`, and the corresponding outcome components are designated using the argument `treats`. For instance, to perform a meta-regression analysis assessing effect modification of SBP on the odds ratio for treatment 3 versus 1, the following command can be applied. Both `z` and `treats` can be specified as vectors to allow for the inclusion of multiple covariates and outcomes.

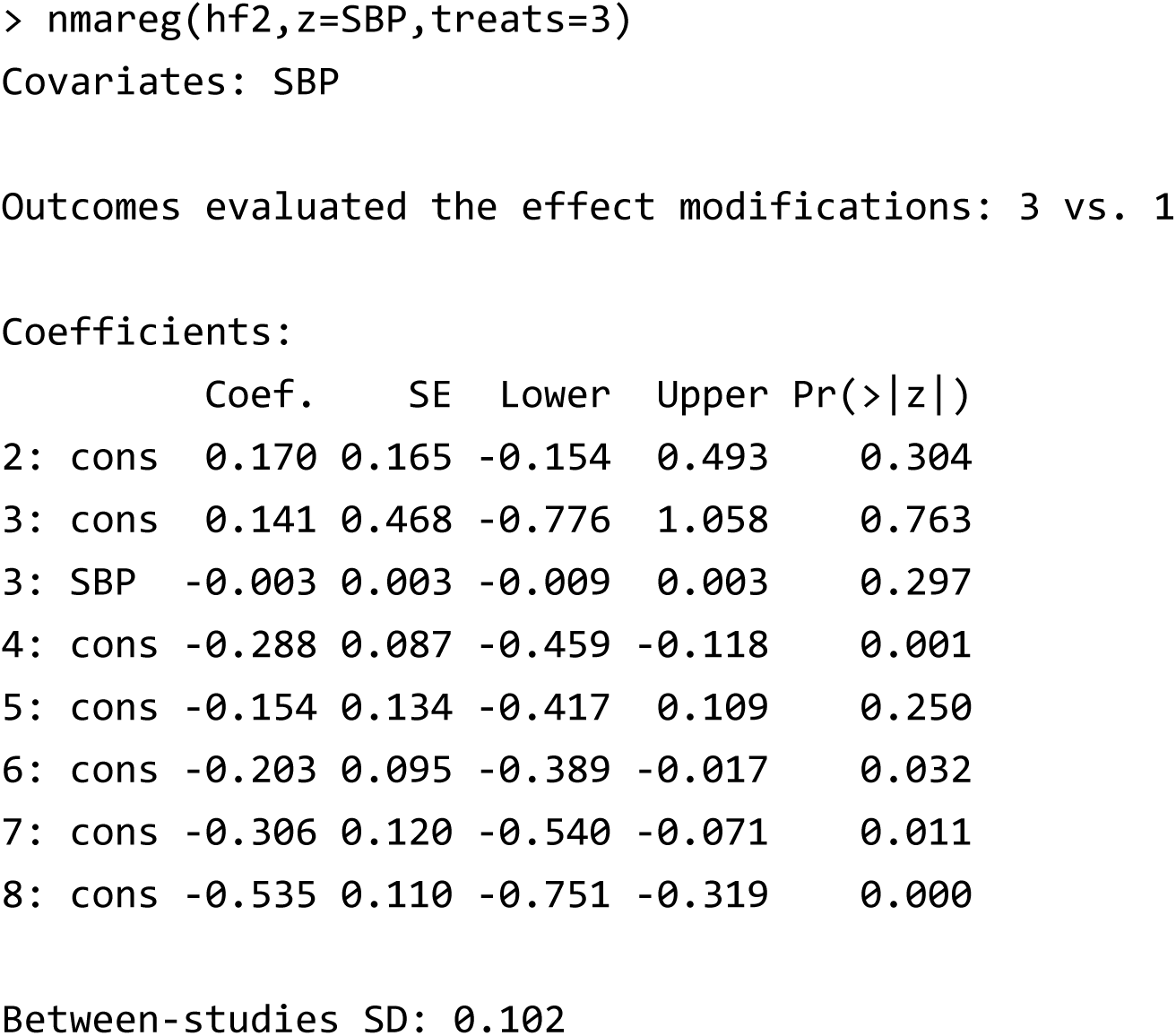

In addition, to estimate the coefficients of each regression function, a sufficient number of direct comparison trials are required. As in ordinary least squares regression, the number of direct comparisons between the corresponding treatments must exceed the number of explanatory variables. If this condition is not met, the estimates from the meta-regression become indeterminate and an error will be produced.

## 6. Analyses of ranking and comparative effectiveness

### 6.1 Ranking probability and SUCRA

As an analytic approach to comparative effectiveness, treatment ranking methods are widely used ^22^. Based on the estimates from model (*), the probabilities of each treatment being ranked best, second, …, or worst can be estimated using a parametric bootstrap procedure ^43^. In addition, the bootstrap method can be used to estimate the surface under the cumulative ranking curve (SUCRA) ^44^. SUCRA is a continuous score ranging from 0 to 1 that indicates the relative position of each treatment within the overall distribution (analogous to a quantile of the distribution). These quantities can be computed using the `nmarank` function, which also provides plots of the ranking probabilities.

> nmarank(hf2) SUCRA:

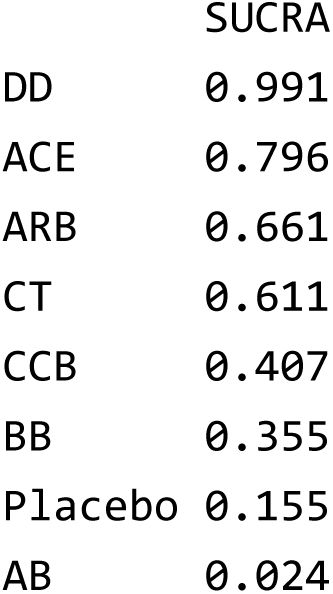

*(The ranking probabilities of each treatment are presented in Figure 2.)*

**Figure 2.**
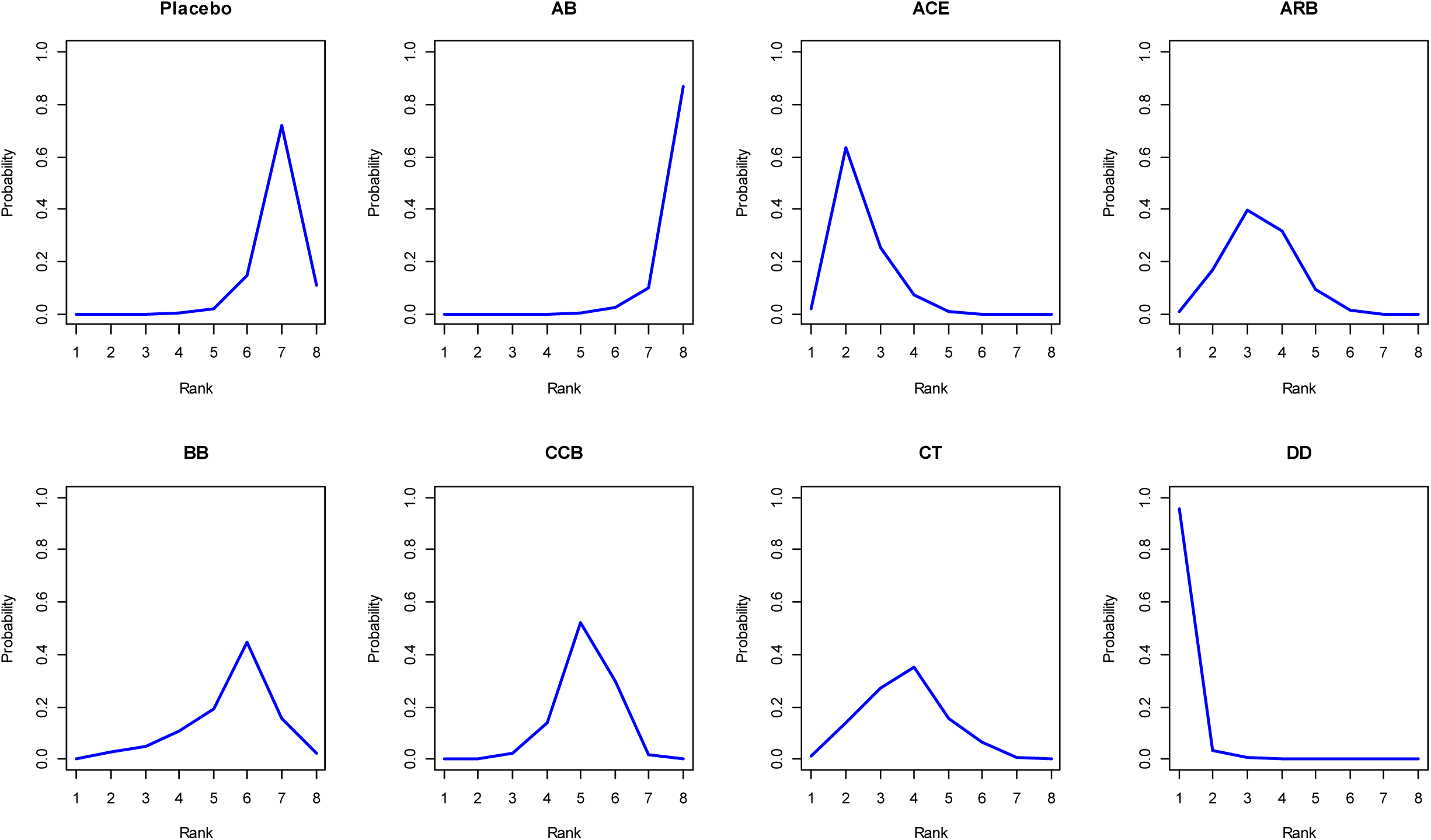
Ranking probability plots for the antihypertensive drug study created by the `nmarank` function.

### 6.2 League table

As another type of output for comparative effectiveness, a league table ^44^ can also be generated easily using the `nmaleague` function. The league table can additionally be exported in CSV format.

Based on the results of the synthesis analysis, a league table can be created with user-specified options. The number of decimal places can be set via the argument `digits`, and the output file name for CSV export can be specified using `out.csv`. For example:

> nmaleague(hf2, eform=TRUE, digits=2, out.csv=“nmaleague_out.csv”)

*(The output is omitted here due to space limitations.)*

### 6.3 Ranked forest plot

The results of the synthesis analysis are often presented graphically using a forest plot. For comparative effectiveness analyses, a ranked forest plot ^45^, in which the estimated effect measures are ordered from largest to smallest, is frequently employed. Such plots can be easily generated using the `nmaforest` function.

> nmaforest(hf2, col.plot=“blue”)

*(The forest plot created is shown in Figure 3.)*

**Figure 3.**
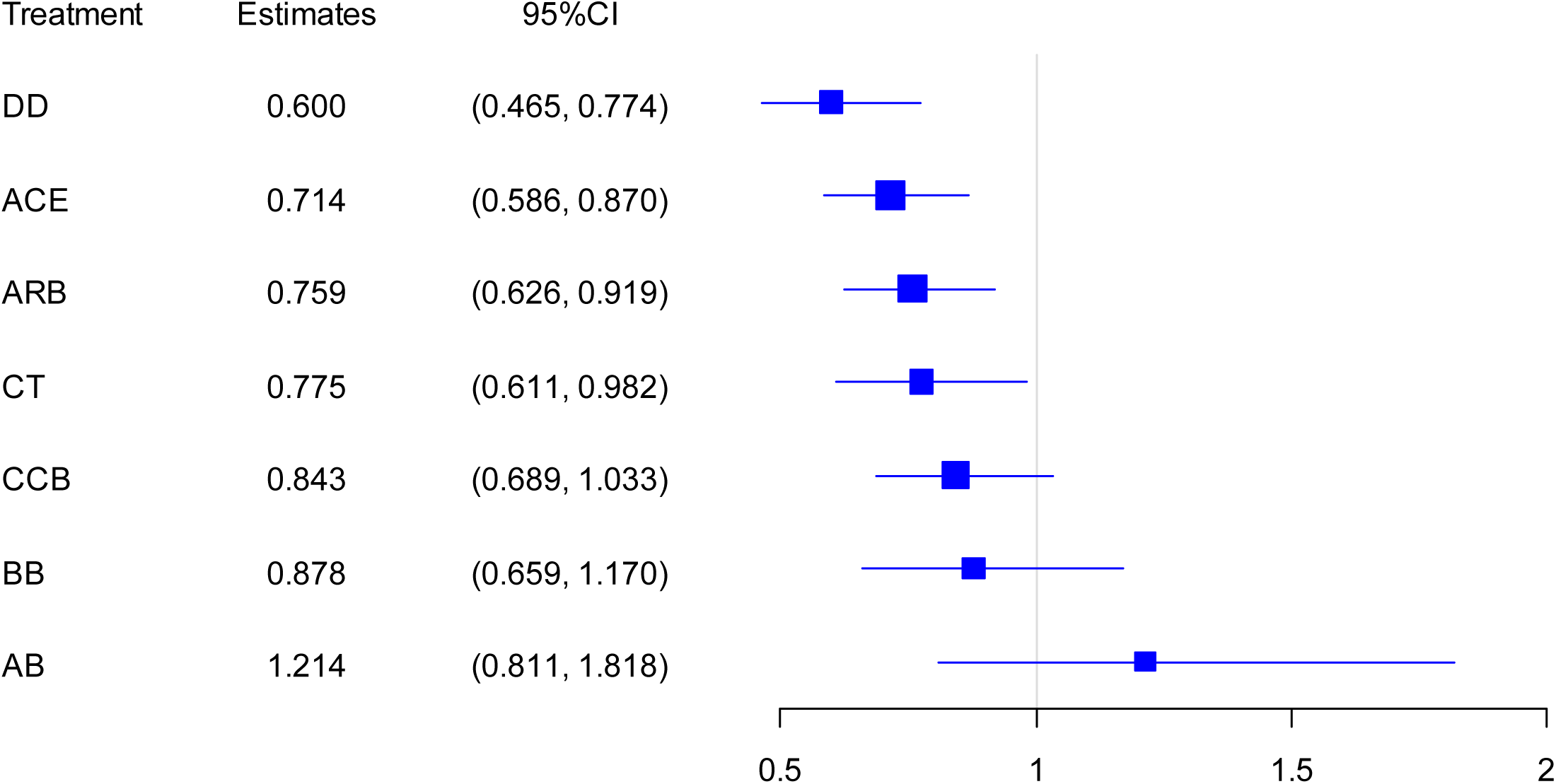
Ranked forest plot for the antihypertensive drug study created by the `nmaforest` function.

## 7. Evaluation of inconsistency

### 7.1 Local inconsistency test

Various statistical methods have been proposed for evaluating inconsistency, and several representative approaches are implemented in the `NMA` package. The first is the local inconsistency test (`local.ict` function). This method extracts triangular loops of three treatments from the evidence network and tests whether the treatment effects obtained from the direct and indirect comparison paths within each loop are consistent ^46,47^.

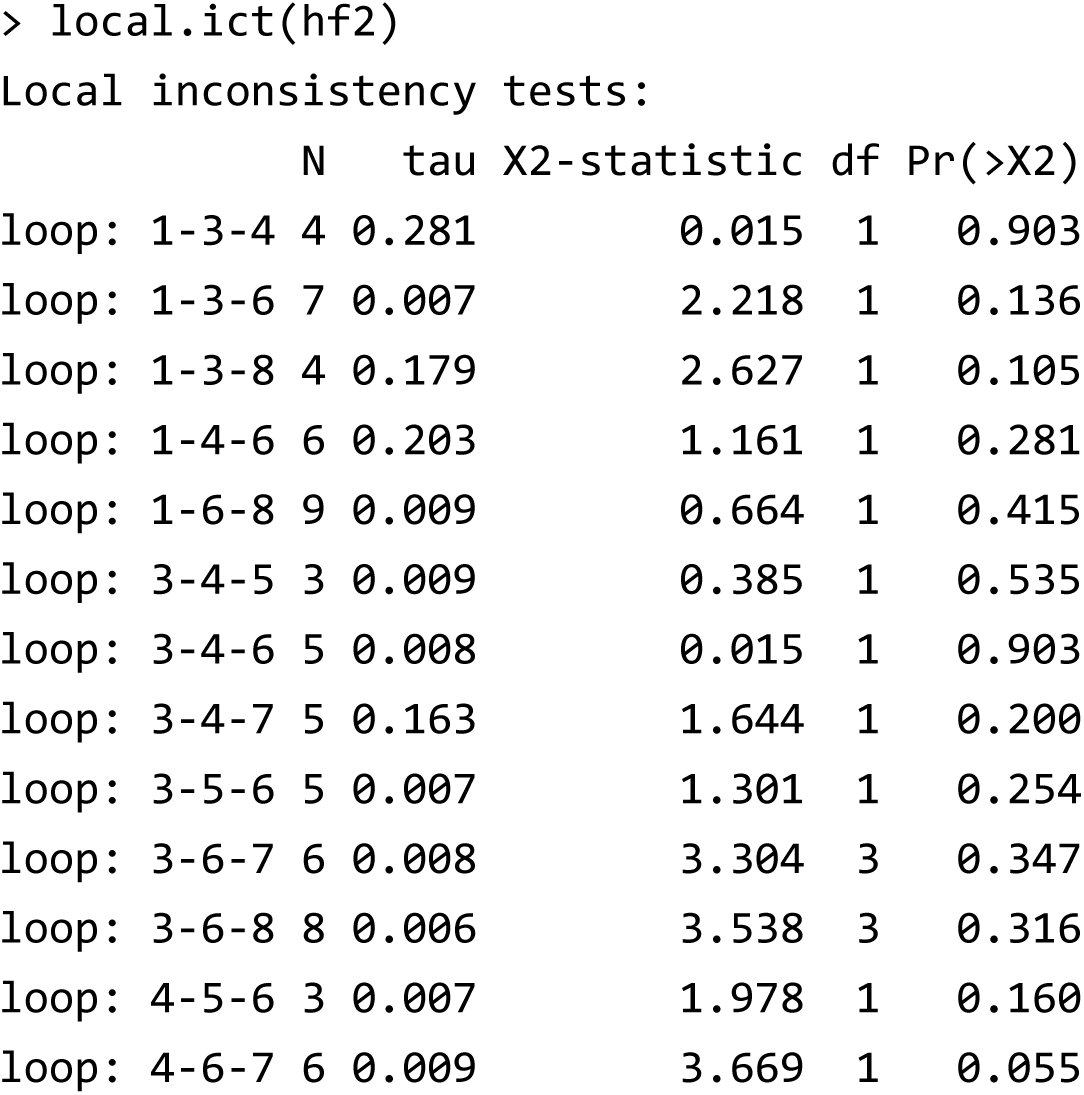

### 7.2 Global inconsistency test

The second method is the global inconsistency test (`global.ict` function). While inconsistency within a triangular loop can be readily understood as the discrepancy between direct and indirect comparisons along the loop, it is less straightforward to define inconsistency across more complex networks. Higgins et al. ^12^ formalized this by defining inconsistency as “differences in treatment effects depending on which treatments are being compared,” and expressed it through a design-by-treatment interaction framework (where “design” refers to the set of treatments being compared) ^48^. In this approach, all possible design-by-treatment interactions in the network are modeled within a meta-regression framework, and a test of the global null hypothesis regarding these interactions was proposed as the global inconsistency test.

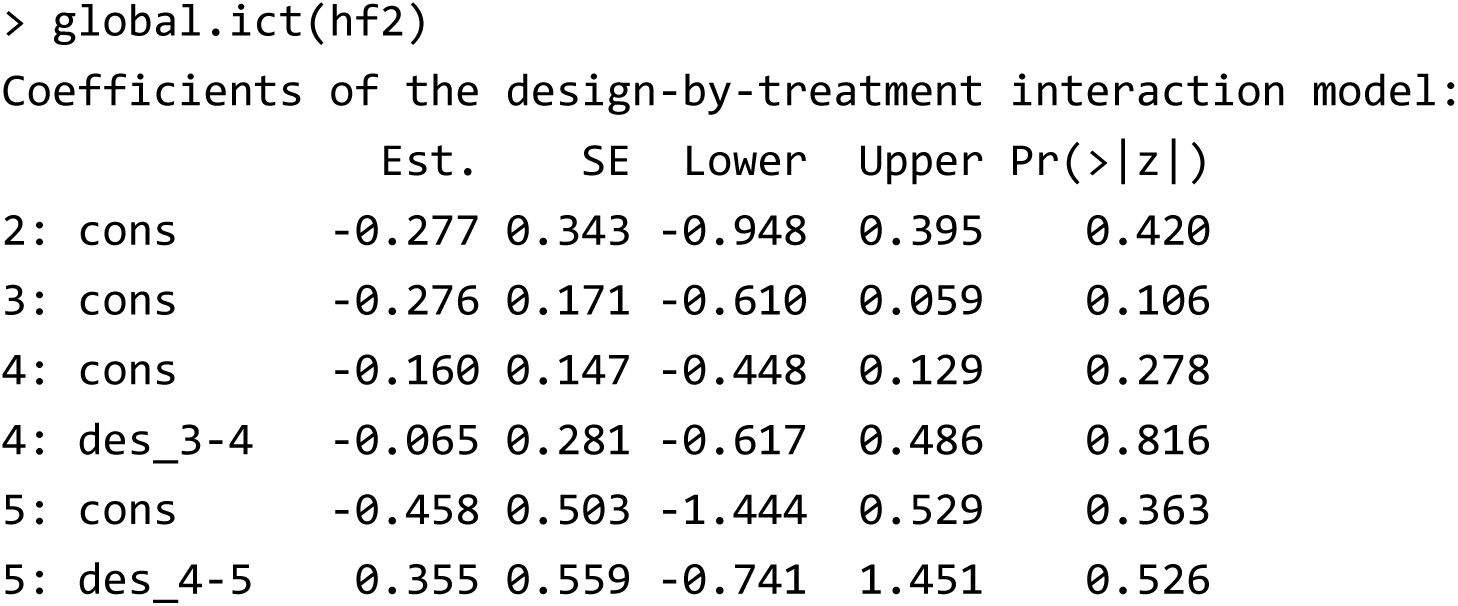

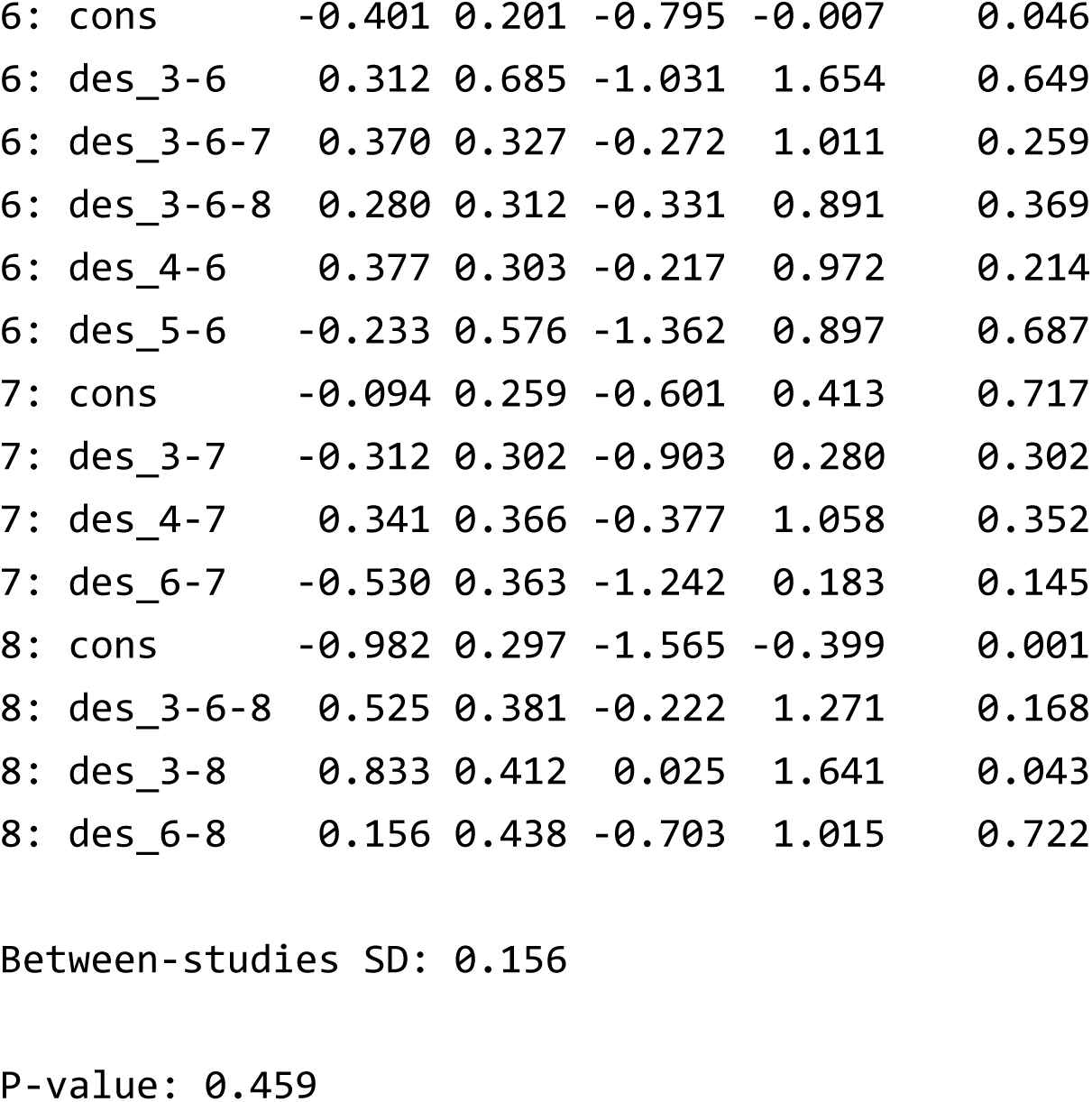

As a supplementary tool, the `nmaQ` function can be used to decompose the *Q*-statistic, providing *Q*-statistics for each design as well as separating the within-design and between-design components. This allows the results of heterogeneity tests to be obtained for each component.

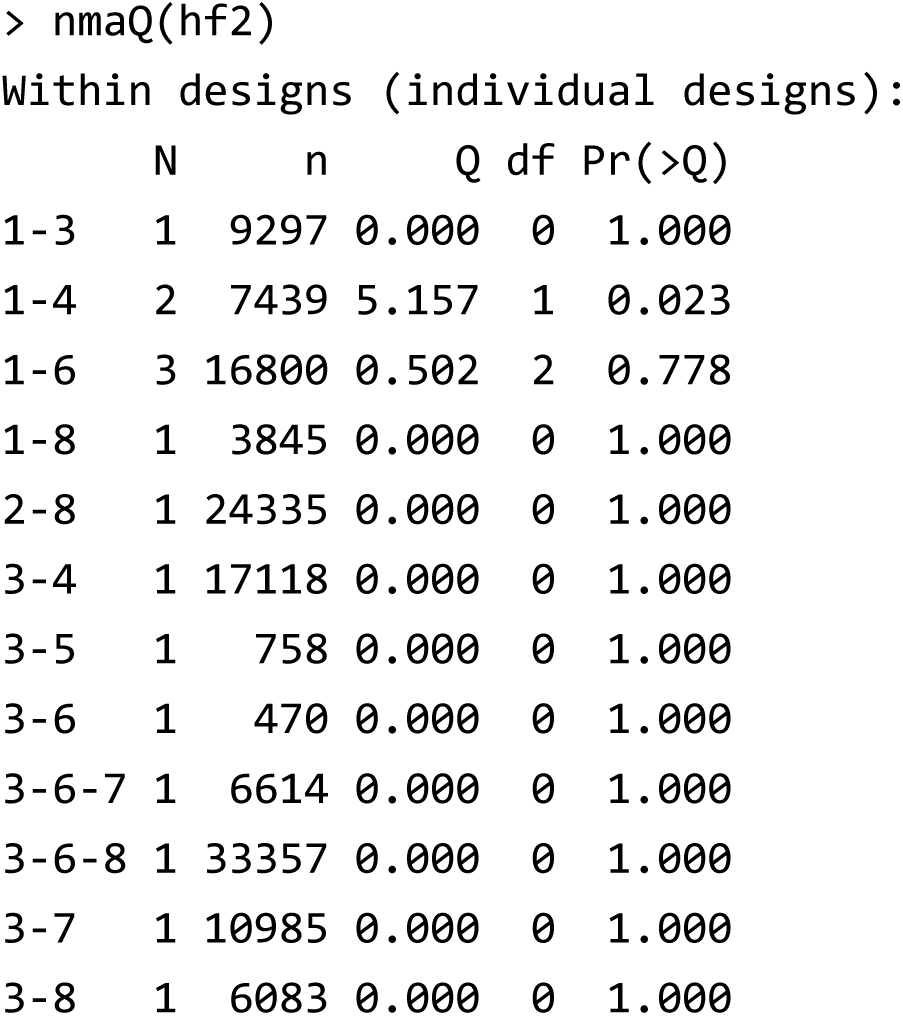

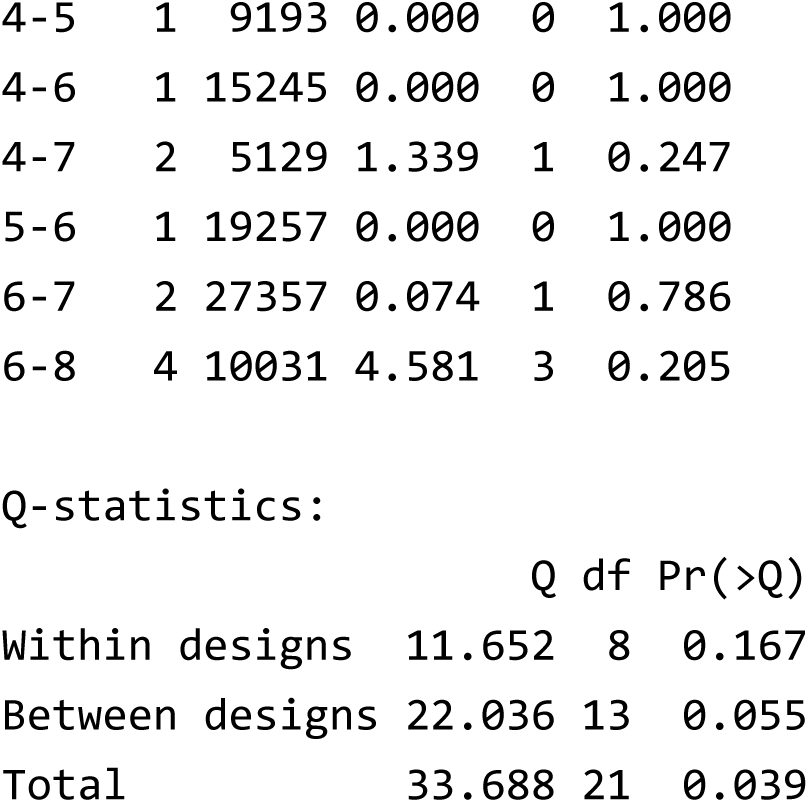

### 7.3 Sidesplitting

The third method is side-splitting (`sidesplit` function). This approach summarizes the direct and indirect evidence for a specific pair of treatments and tests whether their pooled quantities are in disagreement. When multi-arm trials with three or more arms are included, special considerations are required to avoid bias ^49,50^. The `NMA` package adopts the approach proposed by Noma ^49^, which is based on network meta-regression.

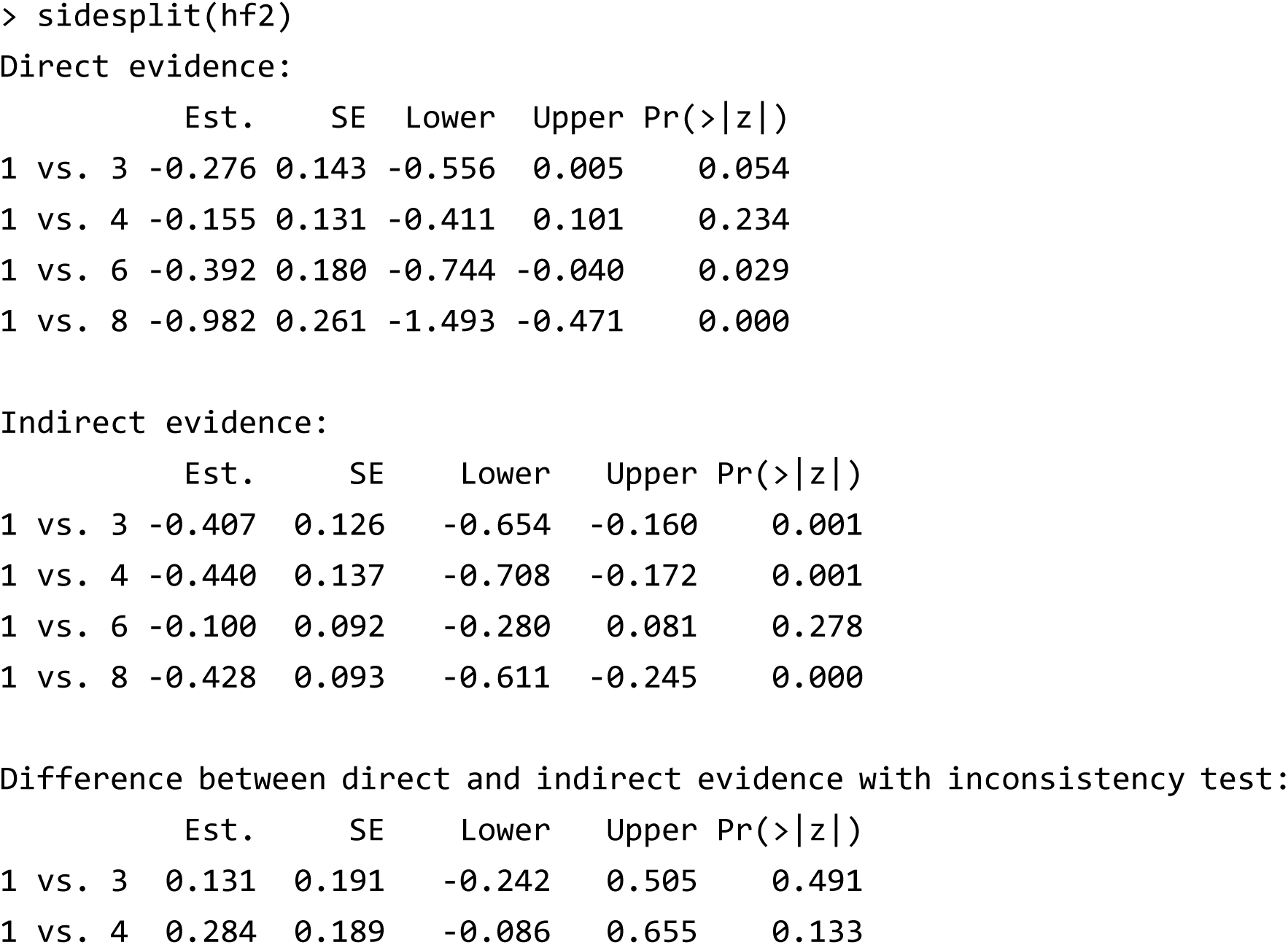

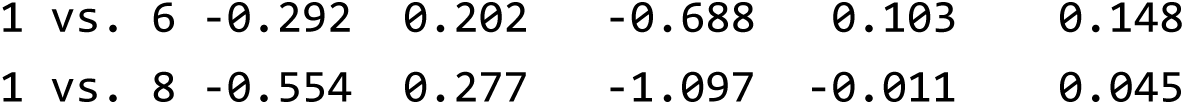

*(Due to space limitations, only the top four rows of results are presented.)*

### 7.4 Random inconsistency model

The fourth method is the random inconsistency model proposed by Jackson et al. ^18^ (`random.icm` function). This approach extends the design-by-treatment interaction meta-regression model of Higgins et al. ^12^ by treating the design effects as random effects ^18,19^. In this model, inconsistency can be evaluated by testing the variance component associated with the random design effects. In addition, this model provides *I*^2^ statistics for heterogeneity, for inconsistency, and for the overall model that jointly accounts for both heterogeneity and inconsistency.

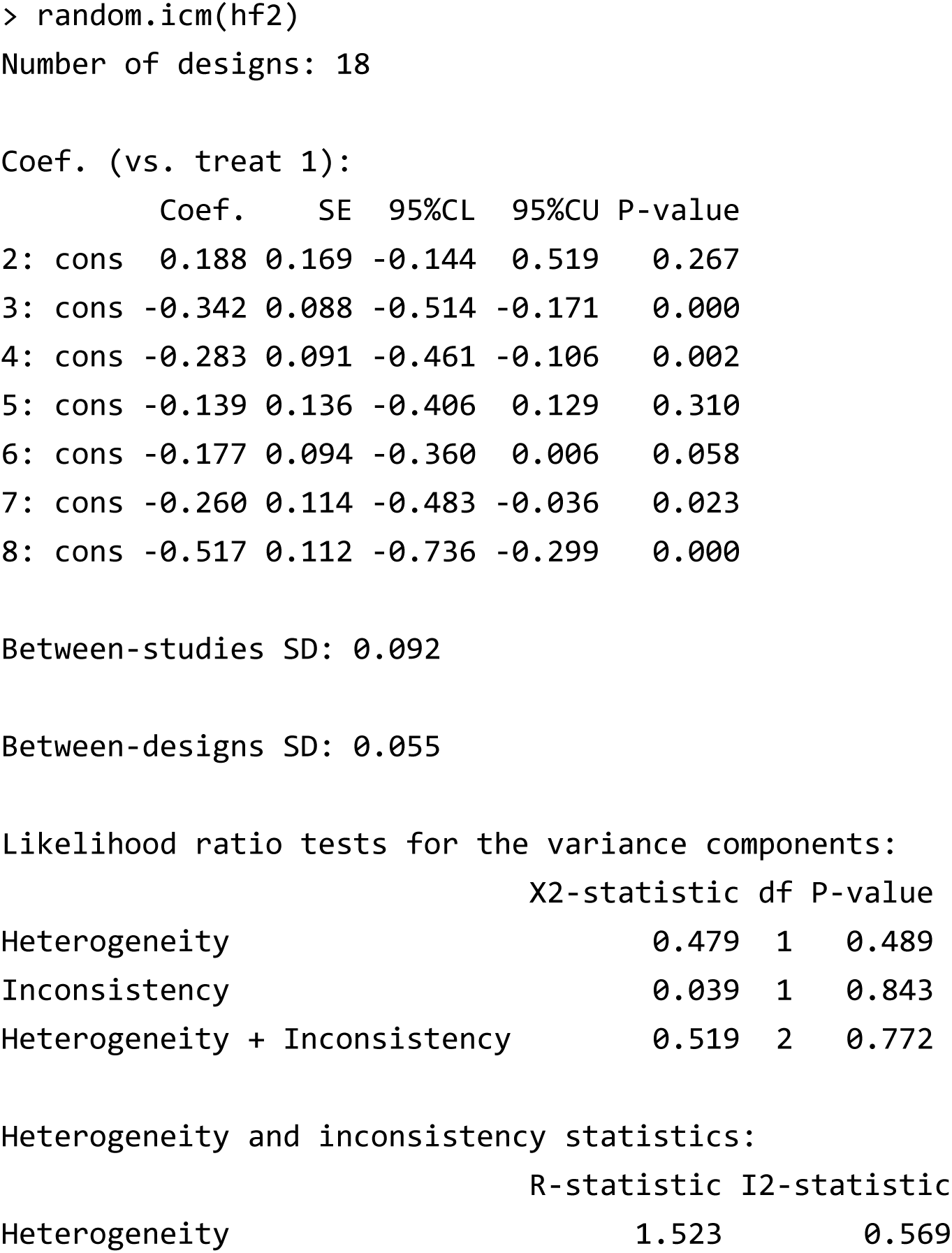

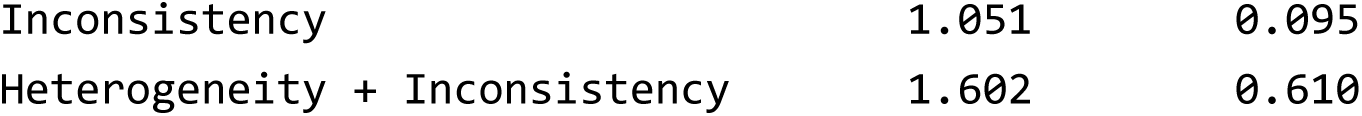

## 8. Other graphical tools

### 8.1 Comparison-adjusted funnel plot

In network meta-analysis, the assessment of publication bias is challenging because multiple treatments are compared and the direction of publication bias may differ across trials. One approach is the comparison-adjusted funnel plot ^43^, in which the results of trials with direct comparisons against a specified reference treatment are adjusted to share a common center line, thereby allowing a descriptive evaluation of publication bias across the network. Such plots can be generated using the `nmafunnel` function.

> nmafunnel(hf2,legends=“bottomright”)

*(The comparison-adjusted funnel plot created is shown in Figure 4.)*

**Figure 4.**
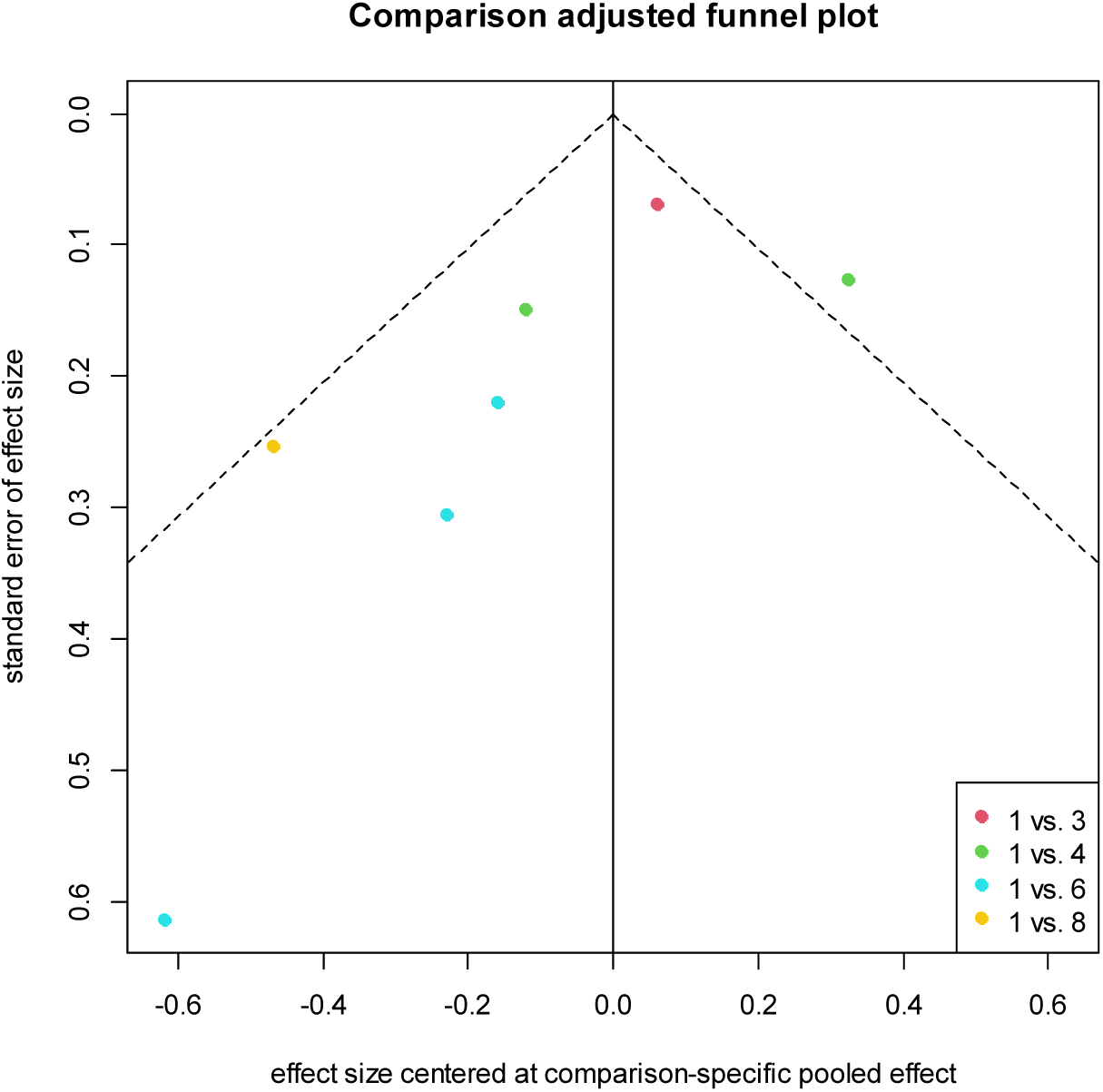
Comparison-adjusted funnel plot for the antihypertensive drug study created by the `nmafunnel` function.

### 8.2 Study weights and contribution matrix

In network meta-analysis, it is important to assess how individual studies influence the synthesized results. Jackson et al. ^51^ and Noma et al. ^50^ demonstrated that contribution rates can be estimated using factorized information, and that contribution weight matrices can be derived accordingly. In the `NMA` package, the `nmaweight` function allows users to compute these contribution weight matrices and to visualize them graphically as a heatmap.

> nmaweight(hf2)

(*The heat plot created is shown in Figure 5.*)

**Figure 5.**
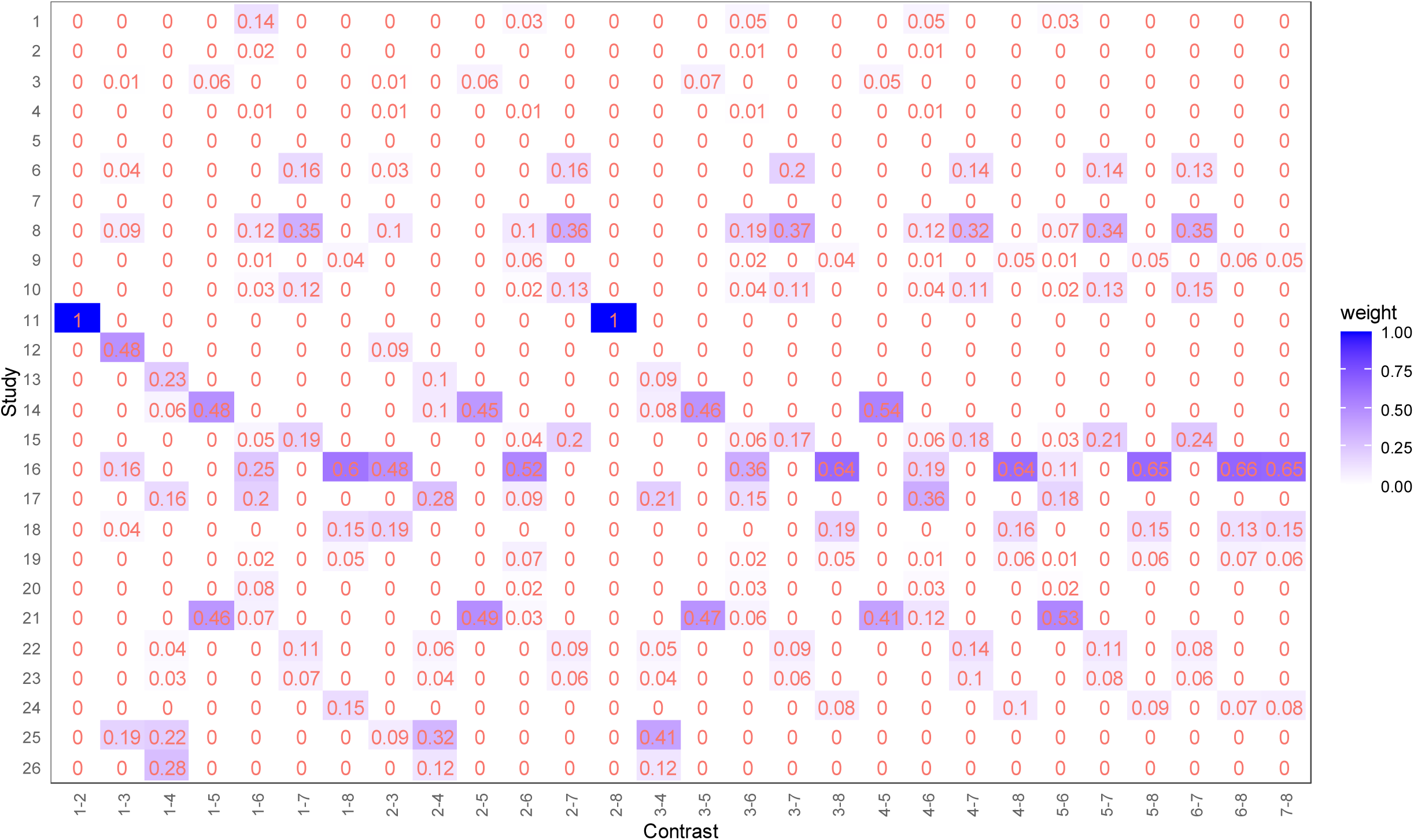
Heat plot for the contribution weights of individual studies for the antihypertensive drug study created by the `nmaweight` function.

### 8.3 Check of transitivity

In network meta-analysis, not only consistency but also transitivity is regarded as a fundamental and essential assumption ^22^, as it underpins the premise that indirect comparisons are theoretically valid even in the absence of direct evidence. To date, no standard statistical method has been firmly established for evaluating transitivity. However, Cipriani, Furukawa, Salanti et al. ^52^ illustrated an approach in which the distributions of key covariates are compared across designs using side-by-side boxplots, thereby enabling an assessment of heterogeneity in covariate distributions. In the `NMA` package, the `transitivity` function generates such side-by-side boxplots to facilitate the evaluation of transitivity.

> transitivity(hf2, SBP, yrange=c(100,220))

(*The side-by-side boxplot created is shown in Figure 6.*)

**Figure 6.**
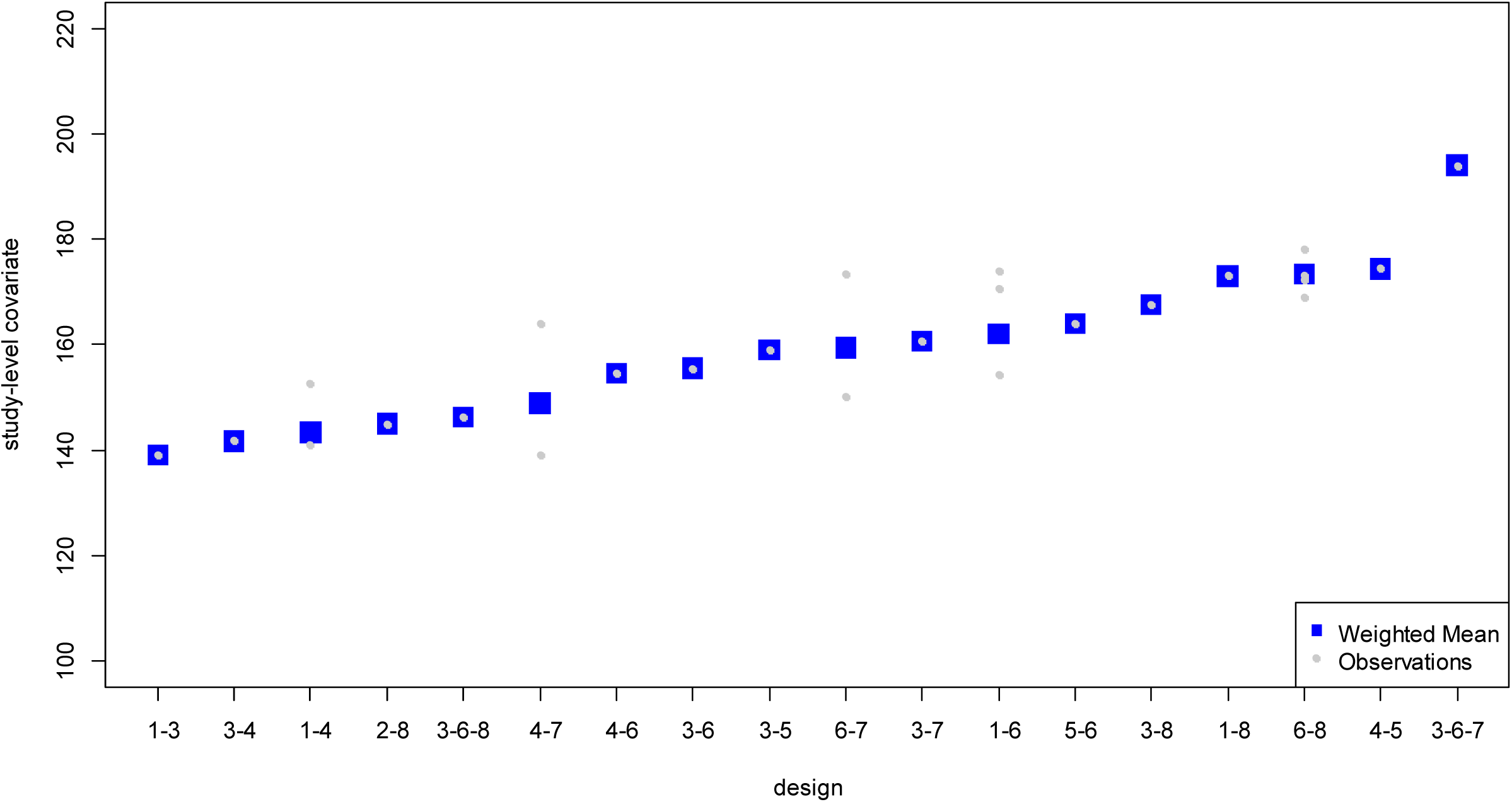
Side-by-side boxplot for assessing transitivity for the antihypertensive drug study created by the transitivity function.

## 9. Concluding remarks

Network meta-analysis is now a well-established methodology within systematic reviews and has already become widely adopted as a standard approach for comparative effectiveness research ^24,53^. However, the statistical methods are often highly technical for non-statisticians. In the practice of systematic reviews, statistical experts are not always involved, and many researchers lack either the time or the technical skills to implement their own programming. Therefore, the availability of user-friendly software with versatile functionality is of critical importance for enabling practitioners to conduct research on important clinical and public health questions. While Graphical User Interface systems such as `NMAstudio` ^54^ and `MetaInsight` ^55^ have been developed to facilitate network meta-analysis, the `NMA` package offers distinct advantages by allowing for flexible, customized analyses within the R environment. Furthermore, it implements more extensive functionalities, including higher-order asymptotic approximations. We hope that the `NMA` package will encourage more researchers and practitioners to undertake network meta-analysis and will contribute to the generation of new evidence that can ultimately improve clinical practice and public health.

## Acknowledgements

This `NMA` package was originally developed for practical training in “Meta-analysis” class at the Kyoto University School of Public Health. We would like to express our gratitude to all those who contributed to this work.

## Author contributions

Conceptualization: H.N., K.M.; Formal analysis: H.N.; Investigation: H.N., K.M.; Methodology: H.N., K.M., S.T., T.A.F.; Supervision: S.T., T.A.F.; Writing—original draft: H.N.; Writing—review and editing: H.N., K.M., S.T., T.A.F.

## Data availability statement

All datasets used in this article are available in `NMA` package on CRAN (https://doi.org/10.32614/CRAN.package.NMA). Also, complete R programs reproducing all case analyses are available on the authors’ GitHub page (https://github.com/nomahi/NMA).

## e-Appendix A: Illustrative example of continuous outcome case

### A.1 Illustrative example: Network meta-analysis of antidiabetic drugs

In this section, we illustrate the application of network meta-analysis for continuous outcomes using the case study by Phung et al. ^1^ on antidiabetic drugs for type 2 diabetes. Phung et al. ^1^ conducted an network meta-analysis comparing six antidiabetic drug classes [α-glucosidase inhibitors (AGI), dipeptidyl peptidase-4 (DPP-4) inhibitors, glinides, glucagon-like peptide-1 (GLP-1) analogues, sulfonylureas, and thiazolidinediones] with placebo. The outcome of interest was the change in HbA1c. For all included trials, arm-level data were available in terms of the mean outcome (`y`), standard deviation (`sd`), and sample size (`n`). When such data are reported at the arm level, all analyses for continuous outcomes can in principle be conducted. In the `NMA` package, this dataset is provided as `antidiabetic`.

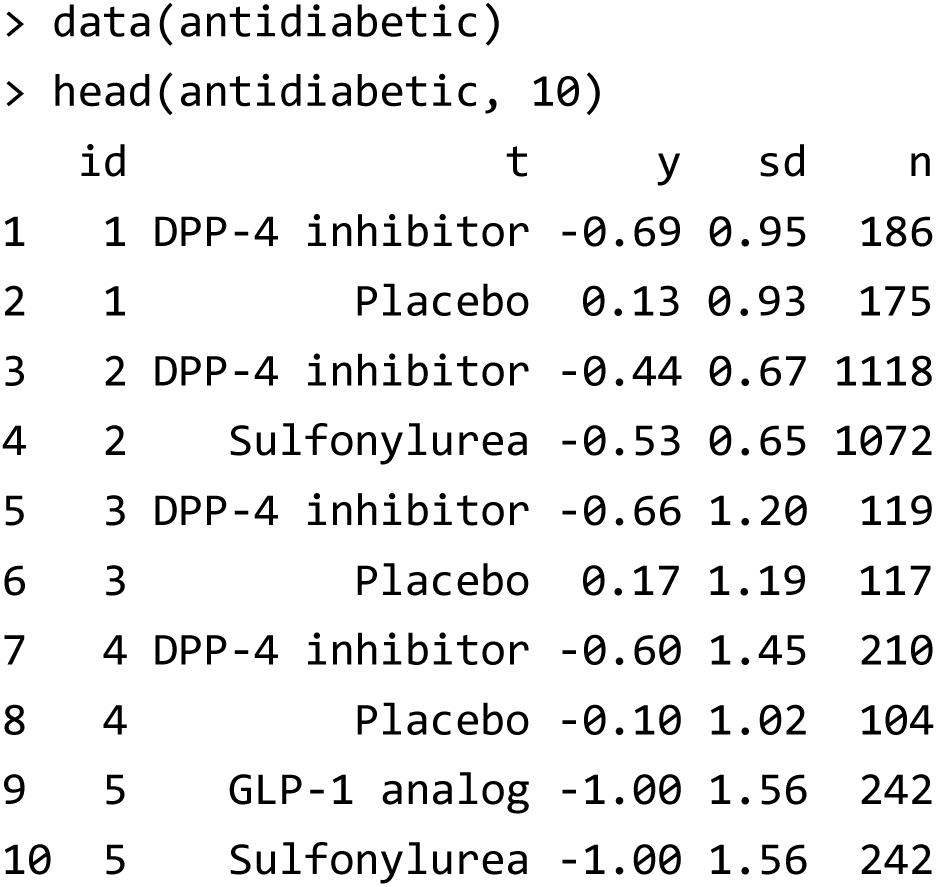

### A.2 Aggregating arm-level and summary level data

On the other hand, in conducting a systematic review, it is not always that arm-level data are reported for all eligible trials. In some studies, only summary statistics, such as an estimated mean difference and its confidence interval, may be available. Such trials cannot be directly incorporated into analyses using the `NMA` package. However, excluding these studies from the analysis would induce a bias analogous to reporting bias.

To address this issue, the `NMA` package provides the function `trans.armdata`, which reconstructs arm-level data from available summary statistics. As an illustration, the dataset `exdataMD` includes summary-level information from three trials in Phung et al. ^1^. When these data are processed with the `trans.armdata` function, the following output is obtained.

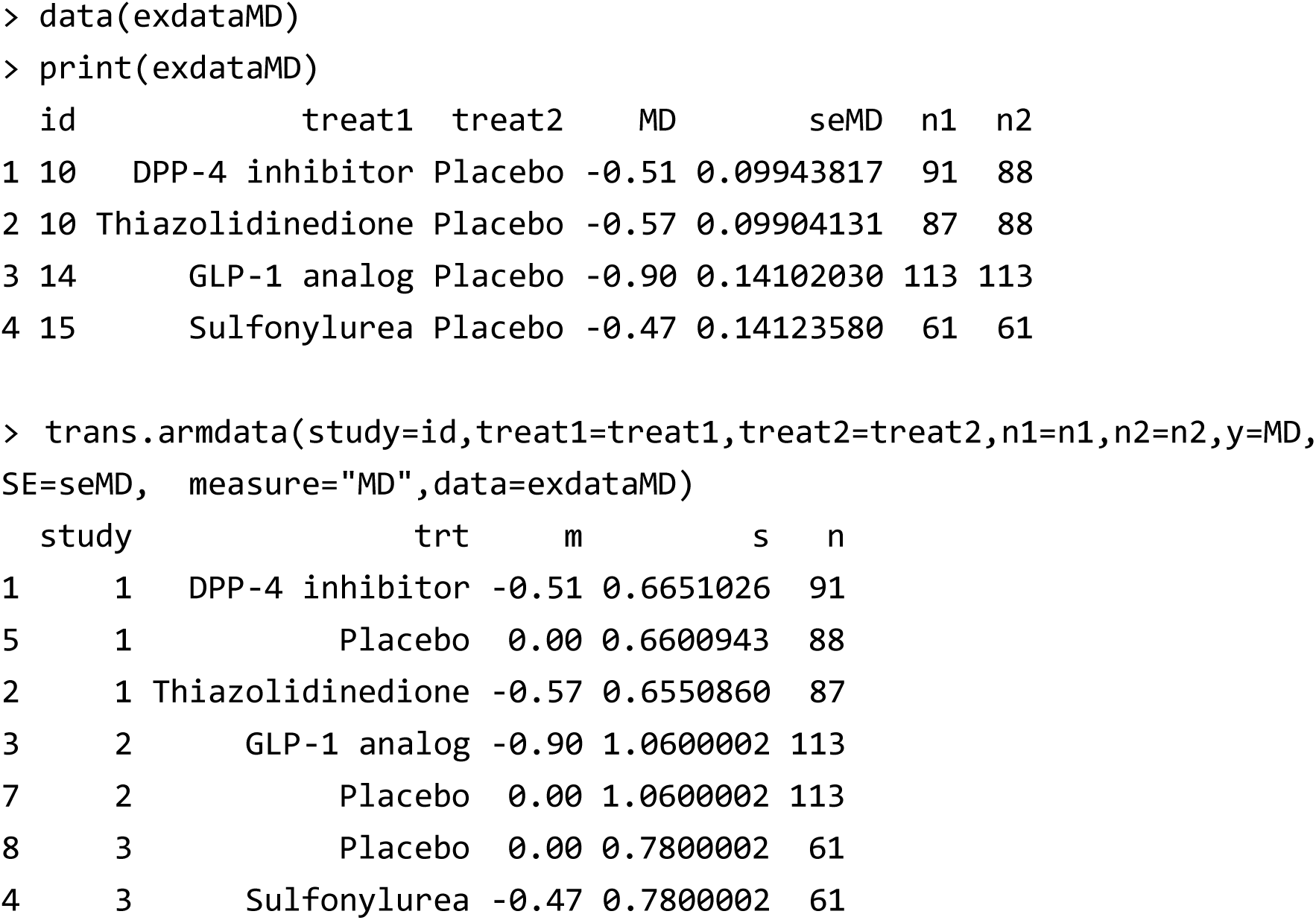

By combining the reconstructed arm-level data with the remaining dataset, it becomes possible to generate an analysis-ready dataset for use in the `NMA` package. An important caveat, however, is that the reconstructed arm-level data may not coincide exactly with the original arm-level data. This discrepancy can arise because the back-calculation is based on large-sample approximations, or because the reported summary statistics were derived from adjusted analyses such as multivariable models.

Nevertheless, since all analyses in the `NMA` package are conducted within a contrast-based framework, the reconstructed arm-level data are subsequently transformed back into summary statistics through the `setup` function prior to analysis. This transformation ensures that the data ultimately used in the network meta-analysis coincide, in principle, with the original reported summary statistics.

### A.3 Data preparation and setup

Once arm-level data have been obtained for all eligible trials—whether directly reported or reconstructed using the methods described in the previous section—the `setup` function can be used to generate a dataset suitable for conducting a contrast-based network meta-analysis. The outcome measure may be expressed either as the mean difference or as the standardized mean difference. In the `NMA` package, the standardized mean difference is, by default, calculated as Cohen’s *d* ^2^.

If summary data on arm-level covariates are available and intended for use in multivariable meta-regression or in assessing transitivity, these variables should be incorporated into the dataset prior to applying the `setup` function. Data editing itself can be conveniently performed using software such as Microsoft Excel before importing the dataset into the `NMA` package.

### A.4 Network meta-analysis

Once the dataset object has been created using the `setup` function, all subsequent analyses can be carried out with simple commands, following the same procedures described in the main text. The illustrative program can be fully reproduced by running R script (B), which is provided at the GitHub page (https://github.com/nomahi/NMA).

As an example, the results of the integrated analysis using the nma function are shown below.

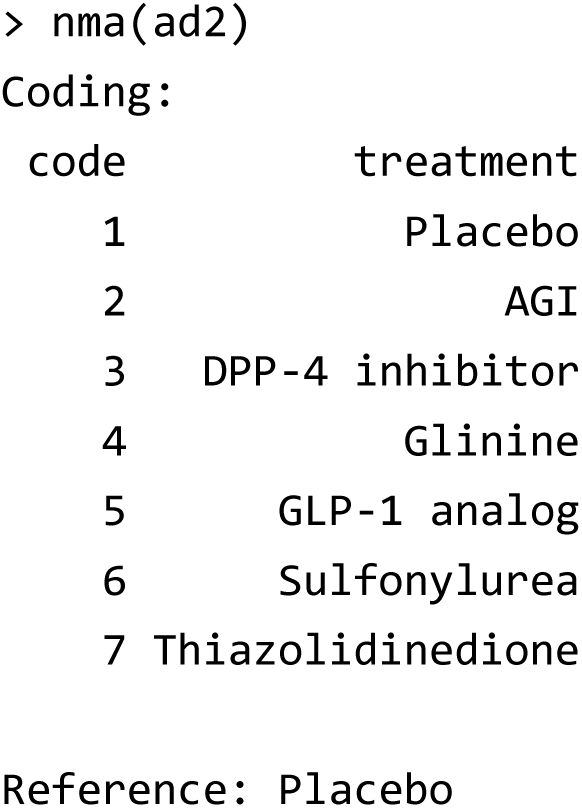

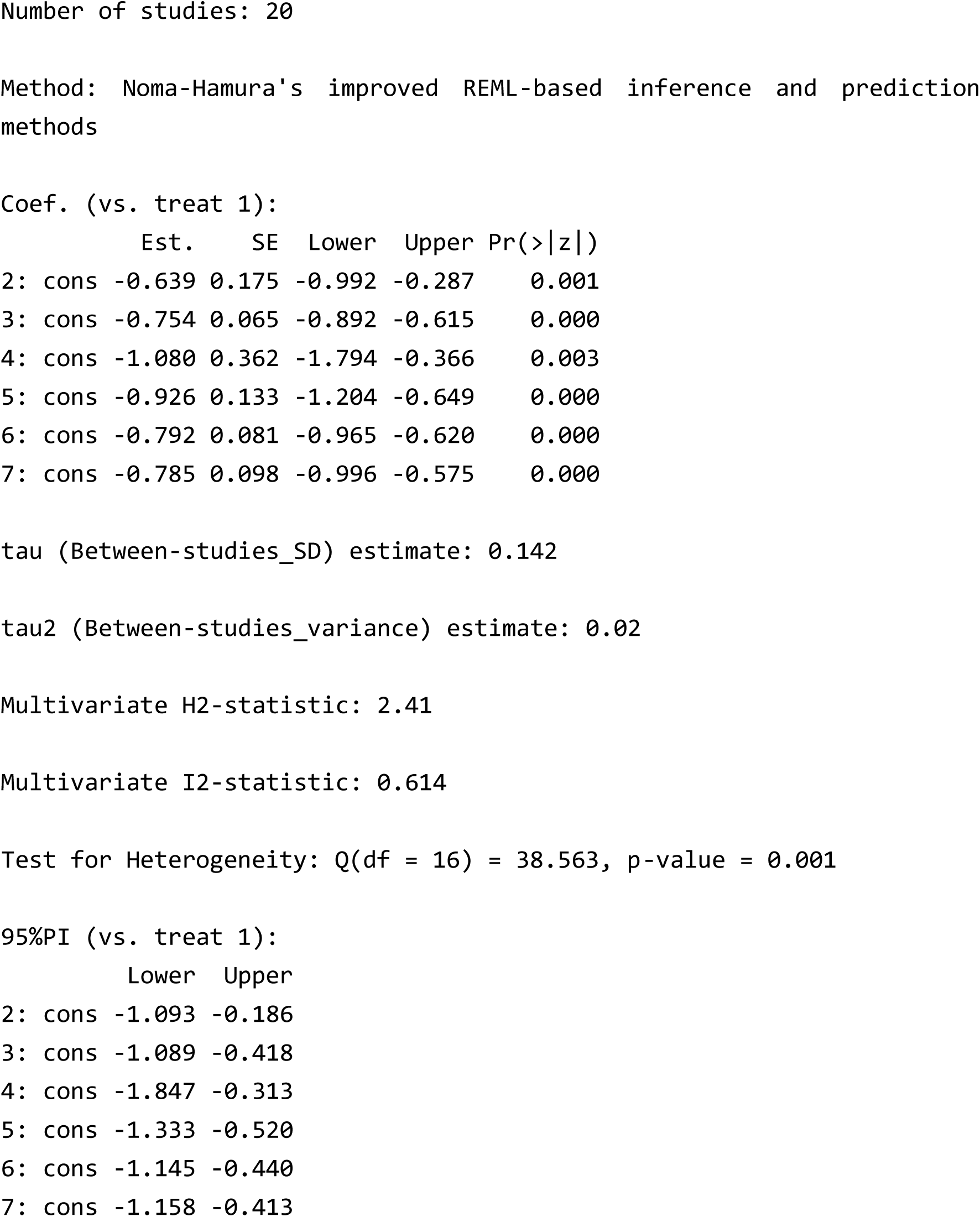

## e-Appendix B: Illustrative example of survival outcome case

### B.1 Illustrative example: Network meta-analysis of COPD

As an example of a survival outcome, we introduce the case study by Woods et al. ^3^, which reported a network meta-analysis of treatments for chronic obstructive pulmonary disease (COPD). In this study, five trials comparing three active treatments—fluticasone, salmeterol, and the salmeterol–fluticasone combination (SFC)—with placebo were synthesized. The outcome of interest was overall survival. However, hazard ratio estimates were available from only two of the trials, whereas in the remaining three trials only the cumulative number of deaths and sample sizes in each arm were reported. These data are provided in the datasets `woods1` and `woods2`.

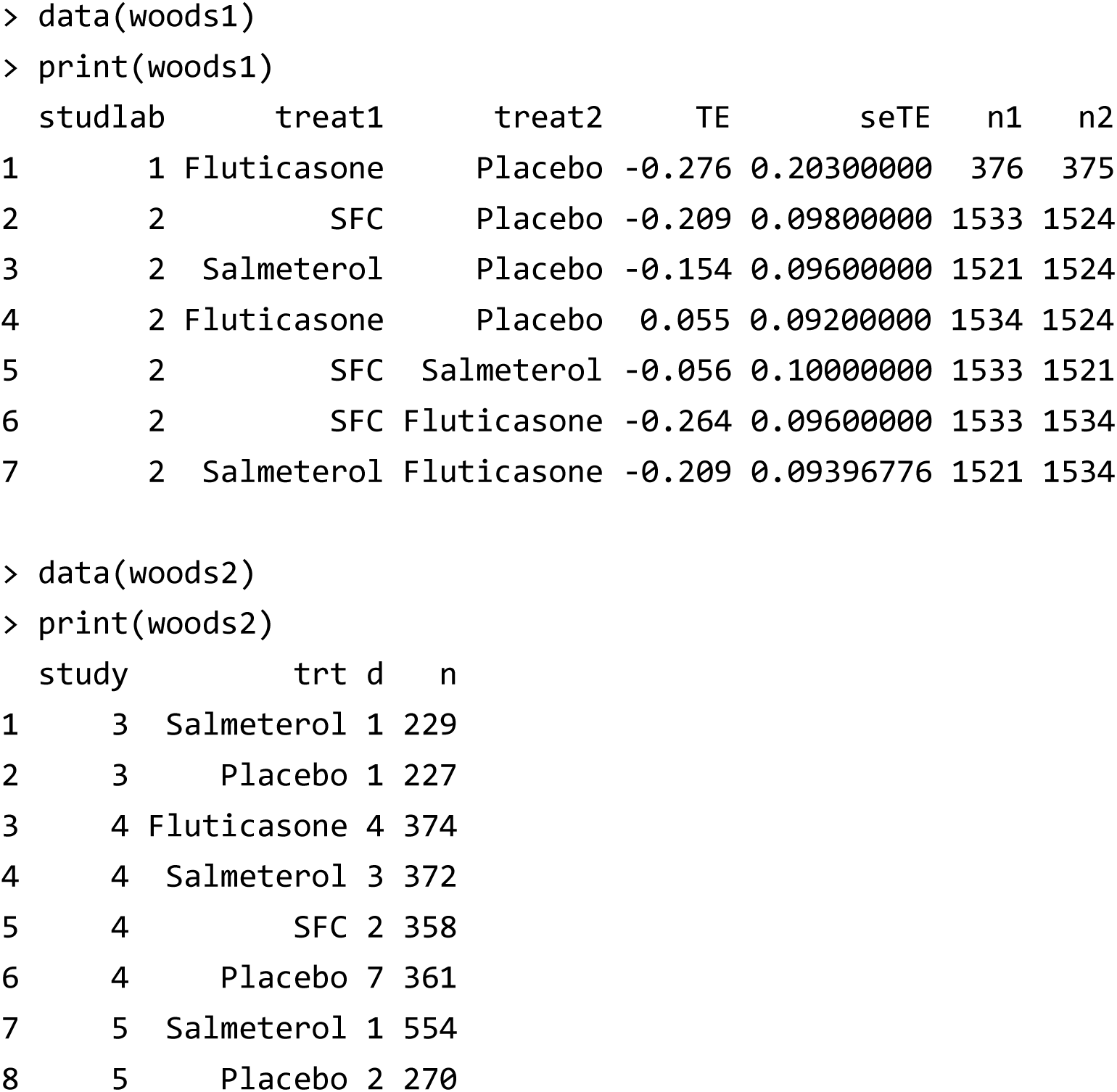

In meta-analyses of time-to-event outcomes, it is common that some trials do not report an estimate of the effect measure of interest—most often the hazard ratio. Parmar et al. ^4^ described methods for deriving hazard ratio estimates from a variety of reported data. In principle, such methods form the basis for reconstructing trial-level information from published reports, and for organizing these into the two types of datasets required as a prerequisite for conducting analyses using the `NMA` package.

### B.2 Aggregating hazard ratio estimates and dichotomized data

For conducting an integrated network meta-analysis, it is necessary to combine these two types of datasets. As noted in the main text, the `NMA` package implements methods based on the contrast-based approach, and thus hazard ratio estimates are synthesized and analyzed using multivariate meta-analysis models. However, covariate summary data—used in meta-regression or transitivity analyses—are often reported at the arm level. To incorporate such covariates into the analysis, it is convenient first to convert the hazard ratio summary statistics into dichotomous data that carry equivalent information (a transformation that can be performed by the `trans.armdata` function).

As in Section A.2, the `trans.armdata` function can be applied to hazard ratio data to reconstruct estimated cumulative numbers of events in each arm, using back-calculation based on the complementary log-log transformation ^5,6^. These reconstructed event counts may not coincide exactly with the original observed counts. Nevertheless, the purpose of these reconstructed data is to serve as “working data” for preprocessing. When passed to the `setup` function, they are transformed back into summary statistics using the same formulas, thereby returning to the original hazard ratio estimates. In this sense, the reconstructed event counts can be regarded as pseudo-data—intermediate quantities created solely for convenience in facilitating subsequent analyses.

When the trans.armdata function is applied to the woods1 dataset, the following arm-level pseudo-data on event counts are obtained.

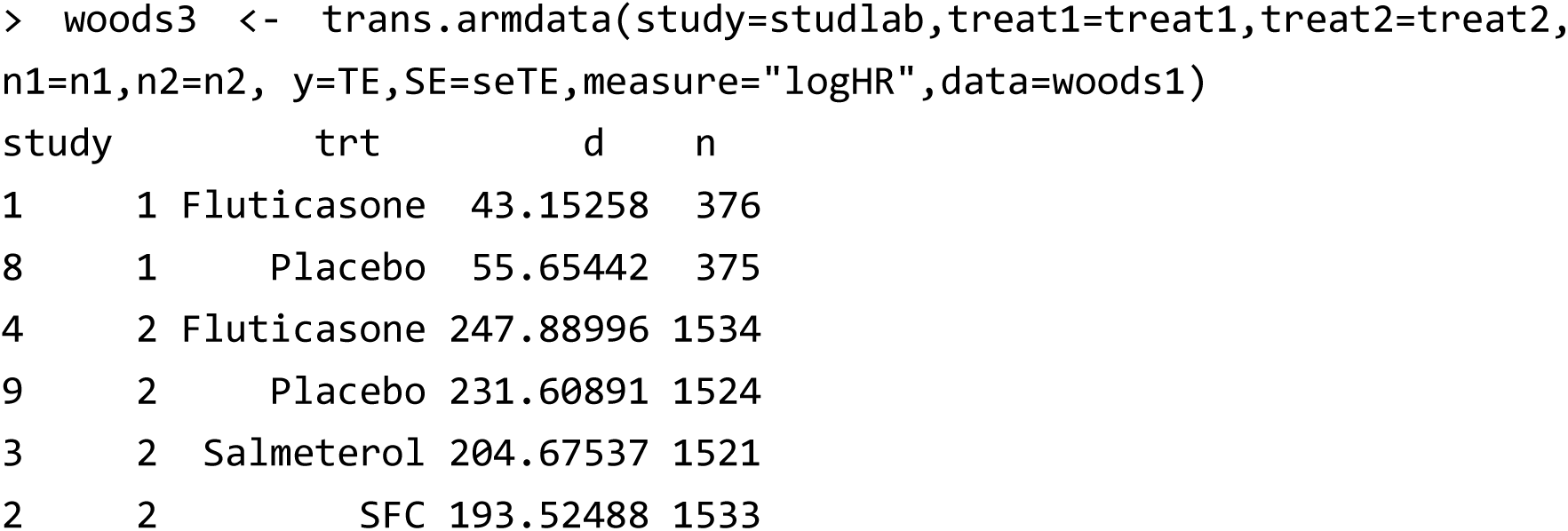

The hazard ratio data are replaced by reconstructed estimates of arm-level event counts. As noted above, these reconstructed values may not exactly match the actual cumulative number of events. However, when processed with the `setup` function, they are transformed back into summary statistics using the same formulas, thereby recovering the original hazard ratio estimates; thus, this does not pose a problem.

By combining the reconstructed arm-level data with the remaining dichotomized datasets, it is then possible to generate a dataset suitable for analysis using the NMA package.

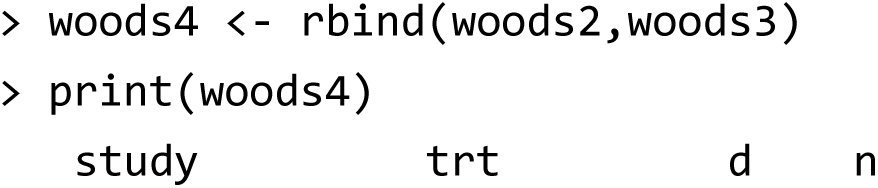

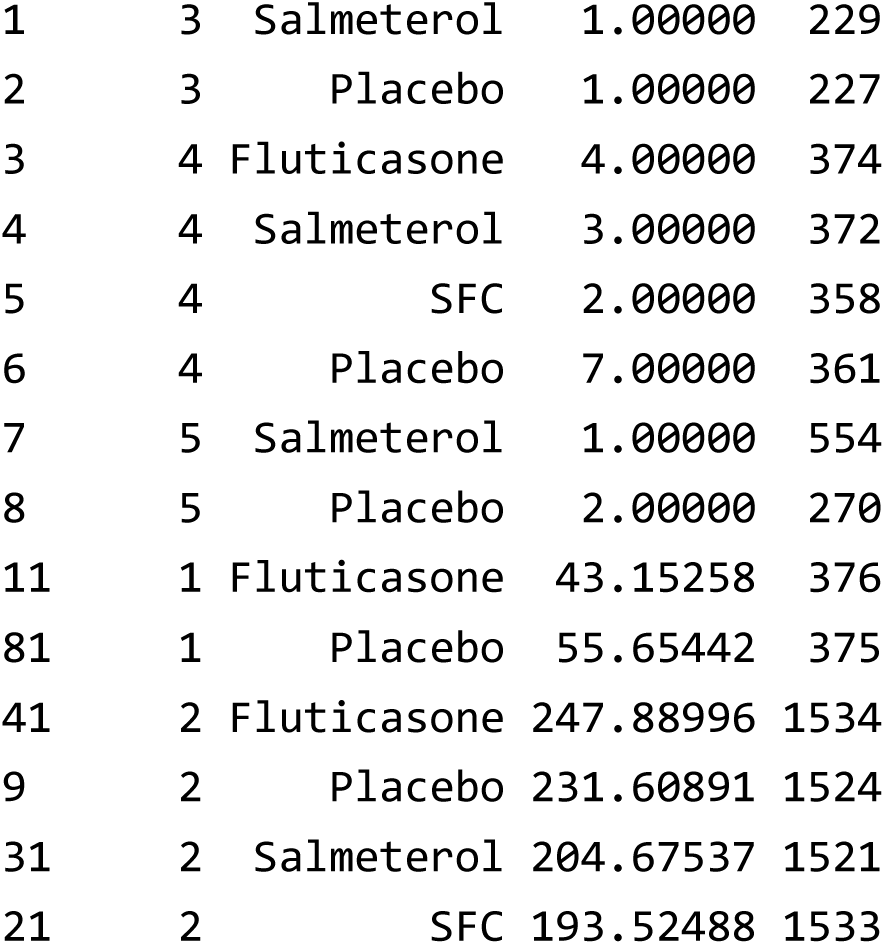

In this way, a single combined dataset can be constructed in the format of event counts and sample sizes. By supplying data in this format to the `setup` function of the `NMA` package, hazard ratio estimates are calculated under the proportional hazards assumption ^5,6^, and a data object is automatically generated for analysis using multivariate meta-analysis models.

If summary data on arm-level covariates are available and intended for use in multivariable meta-regression or transitivity analysis, these variables can simply be added to the dataset object. For practical data handling, the dataset may first be exported as a CSV file and then edited in user-friendly software such as Microsoft Excel before being re-imported for analysis.

### B.3 Data preparation and setup

Using the methods described in the previous sections, once dichotomized arm-level data have been obtained for all eligible trials, the `setup` function can be used to generate a dataset for network meta-analysis based on the contrast-based approach. The outcome measure in this context is the hazard ratio.

When original hazard ratio estimates are available from published reports, the data prior to transformation by the trans.armdata function are, in principle, restored during processing by the setup function.

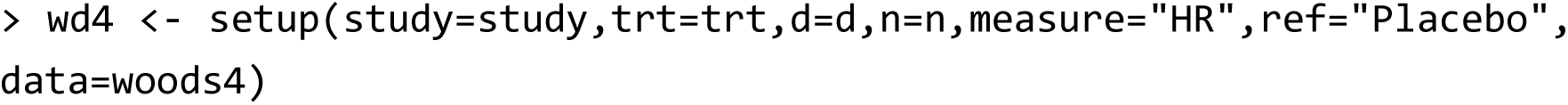

### B.4 Network meta-analysis

Once the hazard ratio dataset object has been created using the `setup` function, all subsequent analyses can be executed with simple commands, following the same procedures as described in the main text. The illustrative program can be fully reproduced by running R script (C), which is provided at the GitHub page (https://github.com/nomahi/NMA).

As an example, the results of the integrated analysis using the nma function are shown below.

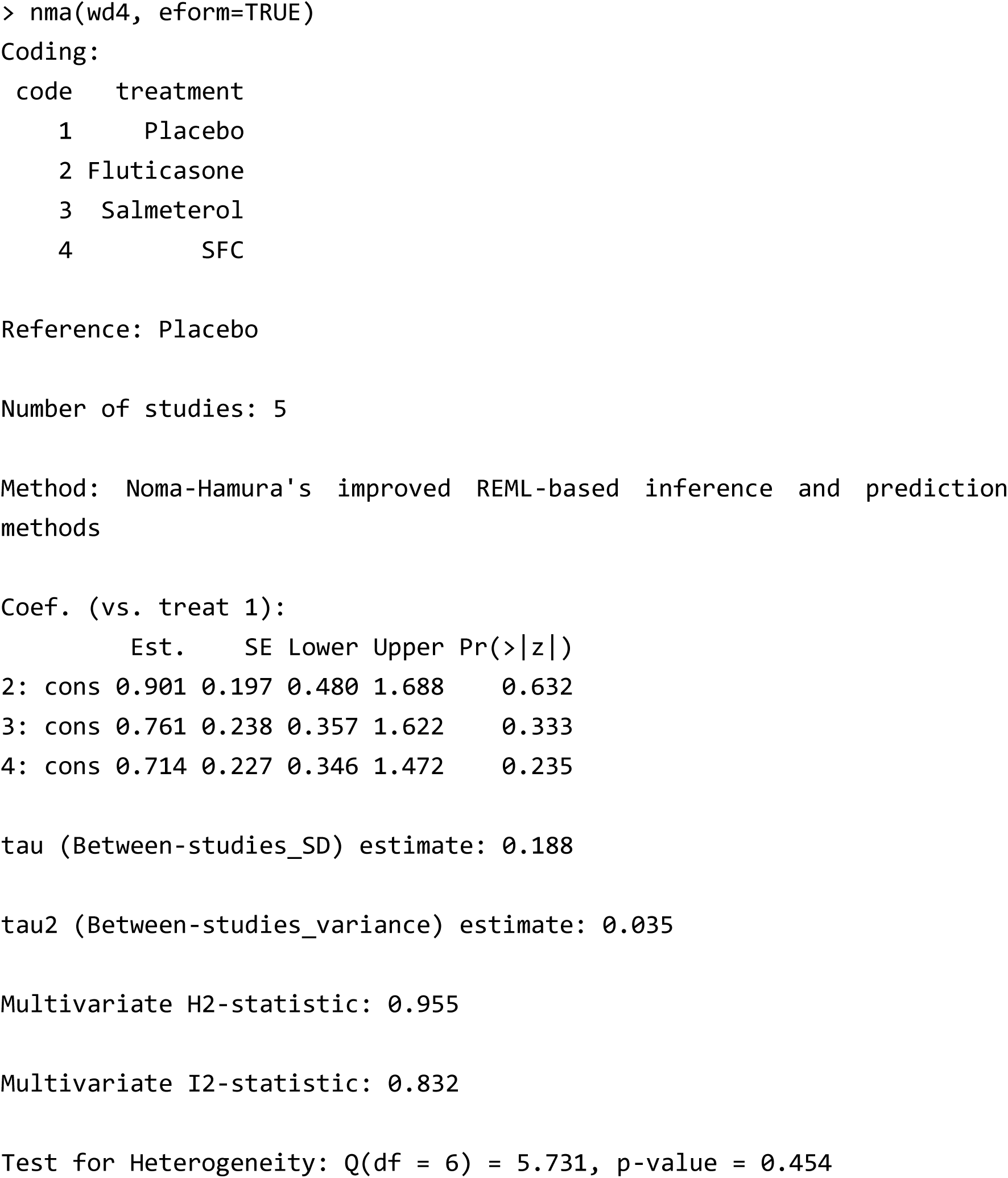

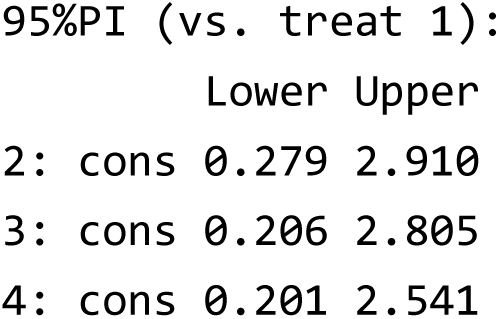

### B.5 Handling the survival probability difference

In systematic reviews of survival outcomes, meta-analyses are sometimes conducted using the difference in survival probabilities as the effect measure ^7,8^, e.g., the difference in 5-year survival rates. Arm-specific event probabilities can be obtained from Kaplan–Meier curves (and their standard errors can be derived, for example, from reported confidence intervals). In network meta-analysis as well, it is possible to use survival probability differences as the effect measure.

The `NMA` package implements a data-processing function, `trans.armdataP`, which converts estimated arm-specific survival probabilities and their standard errors into equivalent information in the form of event counts and sample sizes. These estimates can then be used as working pseudo-data and processed through `setup` in the same manner as hazard ratios, thereby producing contrast-based statistics. Consequently, survival probability differences can be incorporated into all standard `NMA` analyses, alongside other effect measures.

An illustrative example of such a synthesis, using the dataset `exdataP`, is presented below. The illustrative program can be fully reproduced by running R script (D), which is provided at the GitHub page (https://github.com/nomahi/NMA).

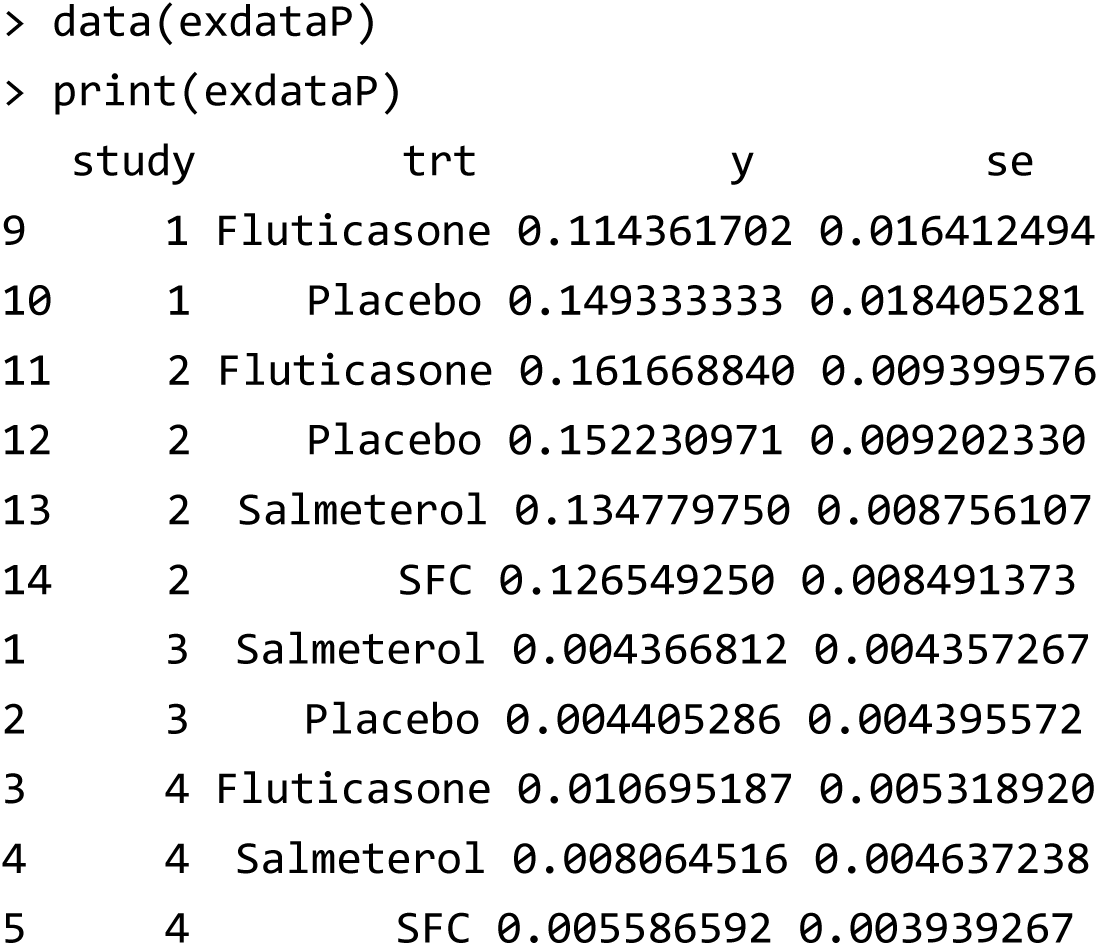

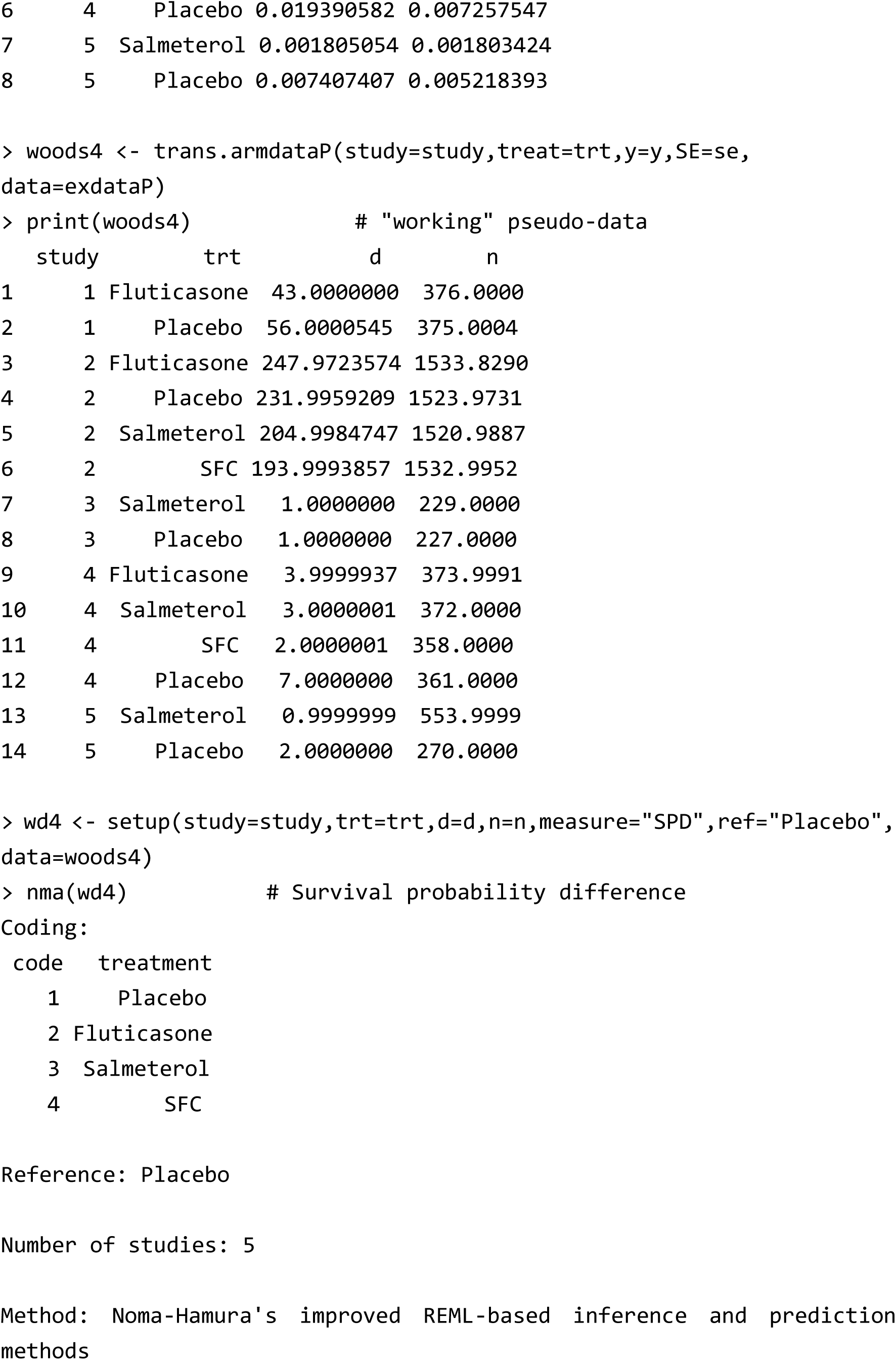

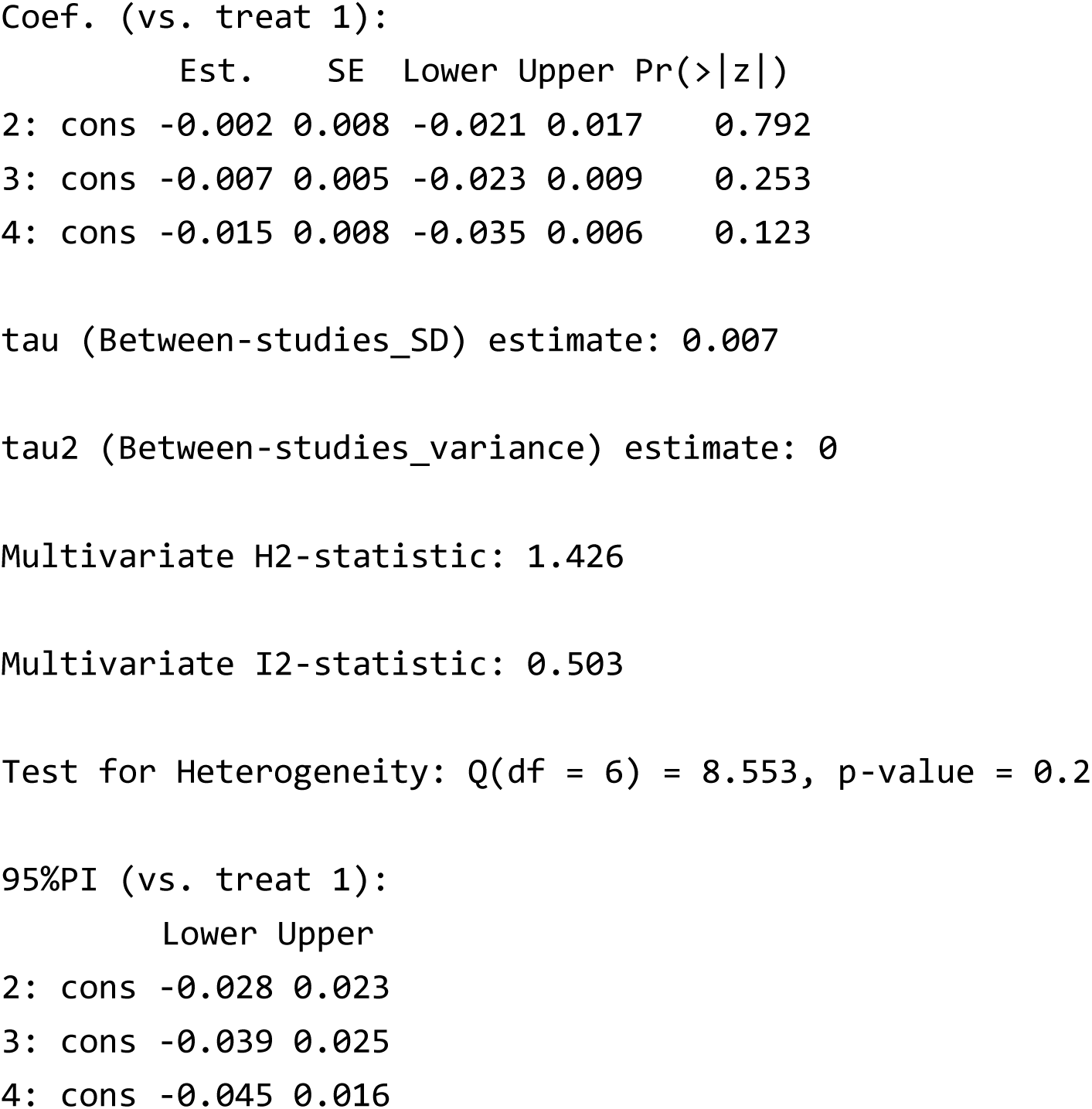

## References

[1] Del Fiol G, Workman TE, Gorman PN. Clinical questions raised by clinicians at the point of care: a systematic review. JAMA Intern Med. 2014;174(5):710–8.

[2] Sox HC, Greenfield S. Comparative effectiveness research: a report from the Institute of Medicine. Ann Intern Med. 2009;151(3):203–5.

[3] Caldwell DM, Ades AE, Higgins JP. Simultaneous comparison of multiple treatments: combining direct and indirect evidence. BMJ. 2005;331:897–900.

[4] Cipriani A, Higgins JP, Geddes JR, Salanti G. Conceptual and technical challenges in network meta-analysis. Ann Intern Med. 2013;159(2):130–7.

[5] Higgins JP, Welton NJ. Network meta-analysis: a norm for comparative effectiveness? Lancet. 2015;386(9994):628–30.

[6] Naci H, O’Connor AB. Assessing comparative effectiveness of new drugs before approval using prospective network meta-analyses. J Clin Epidemiol. 2013;66(8):812–6.

[7] van Valkenhoef G, Lu G, de Brock B, Hillege H, Ades AE, Welton NJ. Automating network meta-analysis. Res Synth Methods. 2012;3(4):285–99.

[8] Lin L, Zhang J, Hodges JS, Chu H. Performing arm-based network meta-analysis in R with the pcnetmeta package. J Stat Softw. 2017;80(2017):5.

[9] Phillippo DM, Dias S, Ades AE, et al. Multilevel network meta-regression for population-adjusted treatment comparisons. J R Stat Soc Ser A. 2020;183(3):1189–1210.

[10] Balduzzi S, Rücker G, Nikolakopoulou A, et al. netmeta: An R Package for network meta-analysis using frequentist methods. J Stat Softw. 2023;106:1–40.

[11] Viechtbauer W. Conducting meta-analyses in R with the metafor package. J Stat Softw. 2010;36:Issue 3.

[12] Higgins JP, Jackson D, Barrett JK, Lu G, Ades AE, White IR. Consistency and inconsistency in network meta-analysis: concepts and models for multi-arm studies. Res Synth Methods.. 2012;3(2):98–110.

[13] White IR, Barrett JK, Jackson D, Higgins JP. Consistency and inconsistency in network meta-analysis: model estimation using multivariate meta-regression. Res Synth Methods.. 2012;3(2):111–125.

[14] White IR. Network meta-analysis. Stata J. 2015;15(4):951–985.

[15] IntHout J, Ioannidis JP, Borm GF. The Hartung-Knapp-Sidik-Jonkman method for random effects meta-analysis is straightforward and considerably outperforms the standard DerSimonian-Laird method. BMC Med Res Methodol. 2014;14:25.

[16] Noma H, Nagashima K, Maruo K, Gosho M, Furukawa TA. Bartlett-type corrections and bootstrap adjustments of likelihood-based inference methods for network meta-analysis. Stat Med. 2018;37(7):1178–1190.

[17] Noma H, Hamura Y, Gosho M, Furukawa TA. Kenward-Roger-type corrections for inference methods of network meta-analysis and meta-regression. Res Synth Methods. 2023;14(5):731–741.

[18] Jackson D, Barrett JK, Rice S, White IR, Higgins JP. A design-by-treatment interaction model for network meta-analysis with random inconsistency effects. Stat Med. 2014;33(21):3639–3654.

[19] Law M, Jackson D, Turner R, Rhodes K, Viechtbauer W. Two new methods to fit models for network meta-analysis with random inconsistency effects. BMC Med Res Methodol. 2016;16:87.

[20] Jackson D, White IR, Riley RD. Quantifying the impact of between-study heterogeneity in multivariate meta-analyses. Stat Med. 2012;31(29):3805–3820.

[21] Noma H, Hamura Y, Sugasawa S, Furukawa TA. Improved methods to construct prediction intervals for network meta-analysis. Res Synth Methods. 2023;14(6):794–806.

[22] Salanti G. Indirect and mixed-treatment comparison, network, or multiple-treatments meta-analysis: many names, many benefits, many concerns for the next generation evidence synthesis tool. Res Synth Methods.. 2012;3(2):80–97.

[23] Sciarretta S, Palano F, Tocci G, Baldini R, Volpe M. Antihypertensive treatment and development of heart failure in hypertension: a Bayesian network meta-analysis of studies in patients with hypertension and high cardiovascular risk. Arch Intern Med. 2011;171(5):384–94.

[24] Higgins JPT, Thomas J. Cochrane Handbook for Systematic Reviews of Interventions. 2nd ed. Wiley-Blackwell; 2019.

[25] Parmar MK, Torri V, Stewart L. Extracting summary statistics to perform meta-analyses of the published literature for survival endpoints. Stat Med. 1998;17(24):2815–2834.

[26] Hedeker D, Siddiqui O, Hu FB. Random-effects regression analysis of correlated grouped-time survival data. Stat Methods Med Res. 2000;9(2):161–79.

[27] Salika T, Turner RM, Fisher D, Tierney JF, White IR. Implications of analysing time-to-event outcomes as binary in meta-analysis: empirical evidence from the Cochrane Database of Systematic Reviews. BMC Med Res Methodol. 2022;22(1):73.

[28] Hutton B, Salanti G, Caldwell DM, et al. The PRISMA extension statement for reporting of systematic reviews incorporating network meta-analyses of health care interventions: checklist and explanations. Ann Intern Med. 2015;162(11):777–784.

[29] DerSimonian R, Laird NM. Meta-analysis in clinical trials. Control Clin Trials. 1986;7(3):177–188.

[30] Egger M, Davey Smith G, Schneider M, Minder C. Bias in meta-analysis detected by a simple, graphical test. BMJ. 1997;315(7109):629–34.

[31] Lu G, Ades AE. Combination of direct and indirect evidence in mixed treatment comparisons. Stat Med. 2004;23:3105–3124.

[32] Jackson D, Riley R, White IR. Multivariate meta-analysis: potential and promise. Stat Med. 2011;30(20):2481–98.

[33] Noma H. Within-study covariance estimators for network meta-analysis with contrast-based approach. Jpn J Biometrics. 2024;44(2):119–126.

[34] Higgins JP, Whitehead A. Borrowing strength from external trials in a meta-analysis. Stat Med. 1996;15(24):2733–2749.

[35] Donegan S, Dias S, Tudur-Smith C, Marinho V, Welton NJ. Graphs of study contributions and covariate distributions for network meta-regression. Res Synth Methods. 2018;9(2):243–260.

[36] Partlett C, Riley RD. Random effects meta-analysis: Coverage performance of 95% confidence and prediction intervals following REML estimation. Stat Med. 2017;36(2):301–317.

[37] Hartung J, Knapp G. On tests of the overall treatment effect in meta-analysis with normally distributed responses. Stat Med. 2001;20(12):1771–82.

[38] Sidik K, Jonkman JN. A simple confidence interval for meta-analysis. Stat Med. 2002;21(21):3153–3159.

[39] Cochran WG. The combination of estimates from different experiments. Biometrics. 1954;10:101–129.

[40] Higgins JPT, Thompson SG. Quantifying heterogeneity in a meta-analysis. Stat Med. 2002;21(11):1539–1558.

[41] Higgins JPT, Thompson SG, Spiegelhalter DJ. A re-evaluation of random-effects meta-analysis. J Royal Stat Soc A. 2009;172(1):137–159.

[42] Nagashima K, Noma H, Furukawa TA. Prediction intervals for random-effects meta-analysis: A confidence distribution approach. Stat Methods Med Res. 2019;28(6):1689–1702.

[43] Chaimani A, Higgins JP, Mavridis D, Spyridonos P, Salanti G. Graphical tools for network meta-analysis in STATA. PloS One. 2013;8:e76654.

[44] Salanti G, Ades AE, Ioannidis JP. Graphical methods and numerical summaries for presenting results from multiple-treatment meta-analysis: an overview and tutorial. J Clin Epidemiol. 2011;64(2):163–71.

[45] Nikolakopoulou A, Higgins JPT, Papakonstantinou T, et al. CINeMA: An approach for assessing confidence in the results of a network meta-analysis. PloS Med. 2020;17(4):e1003082.

[46] Veroniki AA, Vasiliadis HS, Higgins JP, Salanti G. Evaluation of inconsistency in networks of interventions. Int J Epidemiol. 2013;42(1):332–45.

[47] Bucher HC, Guyatt GH, Griffith LE, Walter SD. The results of direct and indirect treatment comparisons in meta-analysis of randomized controlled trials. J Clin Epidemiol. 1997;50(6):683–691.

[48] Jackson D, Boddington P, White IR. The design-by-treatment interaction model: a unifying framework for modelling loop inconsistency in network meta-analysis. Res Synth Methods. 2016;7(3):329–32.

[49] Noma H. Sidesplitting using network meta-regression. Jpn J Biometrics. 2024;44(2):107–118.

[50] Noma H, Tanaka S, Matsui S, Cipriani A, Furukawa TA. Quantifying indirect evidence in network meta-analysis. Stat Med. 2017;36(6):917–927.

[51] Jackson D, White IR, Price M, Copas J, Riley RD. Borrowing of strength and study weights in multivariate and network meta-analysis. Stat Methods Med Res. 2017;26(6):2853–2868.

[52] Cipriani A, Furukawa TA, Salanti G, et al. Comparative efficacy and acceptability of 21 antidepressant drugs for the acute treatment of adults with major depressive disorder: a systematic review and network meta-analysis. Lancet. 2018;391(10128):1357–1366.

[53] Egger M, Higgins JP, Smith GD. Systematic Reviews in Health Research: Meta-Analysis in Context. 3rd ed. BMJ Books; 2022.

[54] Yu T, Metelli S, Chaimani A. NMAstudio: a fully interactive web-application for producing and visualising network meta-analyses. Cochrane Colloquium, London, 2023.

[55] Owen RK, Bradbury N, Xin Y, Cooper N, Sutton A. MetaInsight: An interactive web-based tool for analyzing, interrogating, and visualizing network meta-analyses using R-shiny and netmeta. Res Synth Methods. 2019;10(4):569–581.

## References

[1] Phung OJ, Scholle JM, Talwar M, Coleman CI. Effect of noninsulin antidiabetic drugs added to metformin therapy on glycemic control, weight gain, and hypoglycemia in type 2 diabetes. JAMA. 2010;303(14):1410–8.

[2] Cohen J. Statistical Power Analysis for the Behavioral Sciences. 2nd ed. Lawrence Erlbaum Associates; 1988.

[3] Woods BS, Hawkins N, Scott DA. Network meta-analysis on the log-hazard scale, combining count and hazard ratio statistics accounting for multi-arm trials: a tutorial. BMC Med Res Methodol. 2010;10:54.

[4] Parmar MK, Torri V, Stewart L. Extracting summary statistics to perform meta-analyses of the published literature for survival endpoints. Stat Med. 1998;17(24):2815–2834.

[5] Hedeker D, Siddiqui O, Hu FB. Random-effects regression analysis of correlated grouped-time survival data. Stat Methods Med Res. 2000;9(2):161–79.

[6] Salika T, Turner RM, Fisher D, Tierney JF, White IR. Implications of analysing time-to-event outcomes as binary in meta-analysis: empirical evidence from the Cochrane Database of Systematic Reviews. BMC Med Res Methodol. 2022;22(1):73.

[7] Maas C, Kent DM, Dinmohamed AG, van Klaveren D. Quantifying absolute treatment effect heterogeneity for time-to-event outcomes across different risk strata: divergence of conclusions with risk difference and restricted mean survival difference. medRxiv. 2024; DOI: 10.1101/2024.12.19.24319347.

[8] Higgins JPT, Thomas J. Cochrane Handbook for Systematic Reviews of Interventions. 2nd ed. Wiley-Blackwell; 2019.

